# PRESCOTT: a population aware, epistatic and structural model accurately predicts missense effect

**DOI:** 10.1101/2024.02.03.24302219

**Authors:** Mustafa Tekpinar, Laurent David, Thomas Henry, Alessandra Carbone

## Abstract

Predicting the functional impact of point mutations is a complex yet vital task in genomics. PRESCOTT stands at the forefront of this challenge and reconstructs complete mutational landscapes of proteins, enables the identification of protein regions most vulnerable to mutations and assigns scores to individual mutations, assisting pathologists in evaluating the pathogenic potential of missense variants. PRESCOTT categorizes these variants into three distinct classes: Benign, Pathogenic, or Variants of Uncertain Significance (VUS). The model leverages protein sequences across millions of species, advanced protein structural models, and extensive genomic and exomic data from diverse human populations. By using only sequence and structural information, it significantly improves on current standards for predicting mutations in human proteins and matches AlphaMissense performance, which incorporates allele frequency data in its analysis. By including population-specific allele frequencies, PRESCOTT excels in genome-scale score separation of ClinVar benign and pathogenic variants and surpasses AlphaMissense in analyzing the ACMG reference human dataset and the over 1800 proteins from the Human Protein Dataset. Its efficacy is particularly notable in autoinflammatory diseases, accurately predicting pathogenic gain-of-function missense mutations, a task known for its difficulty. Efficiency and accessibility are key aspects of PRESCOTT. The user-friendly PRESCOTT webserver facilitates mutation effect calculations on any protein and protein variants. The server hosts a Comprehensive Human Protein Database for over 19,000 human proteins, based on sequences and structures, ready for a customized allele population analysis. Additionally, the tool provides open access to all intermediate scores, ensuring interpretability and transparency in variant analysis. PRESCOTT is a significant stride forward in the field of genomic medicine, offering unparalleled insights into protein mutational impacts.

## INTRODUCTION

Proteins play a critical role in cellular processes: understanding what proteins do when they interact within complex systems is crucial for unveiling the underlying mechanisms that govern life. Yet, proteins themselves exhibit remarkable complexity, with a single protein potentially having multiple functions depending on their interacting molecules, (alternative) folding, post transcriptional modifications, cellular compartments, or the organisms that it belongs to. Despite advances in experimental and computational methods, only a small fraction of known proteins has been fully functionally characterized (1,2). Moreover, current research on the molecular foundations of life and disease has focused primarily on a limited set of well-known proteins, resulting in a bias in protein annotation (3–5). For the functional characterization of the human proteome, for instance, this bias is massive since there are only 5,000 particularly well-studied human proteins (6) over the ∼20,000 in the full proteome (3,7,8). Thus, given the enormous diversity of proteins and their multiple functions, the understanding of how these proteins recognize other molecules and how sensitive they are to mutations would transform our comprehension of the processes that drive many key biological functions and genetic diseases. Indeed, missense mutations, that is non-synonymous mutations induced by single nucleotide changes, can alter protein function through various mechanisms, such as disrupting protein stability, modulating enzyme activity allosterically, and changing Protein-Protein Interactions (PPIs). Understanding the effects of missense mutations, whether they are pathogenic or benign in the human proteome can help rationalize potential mechanisms of disease-related interface mutations and improve molecular diagnosis. More globally, the identification of the mutations that can affect or not a protein’s action provides a huge amount of information for the design of experiments heading to advances on our comprehension of the functional roles of proteins. Furthermore, pharmaceutical research can be facilitated if protein mutational landscapes can be studied accurately and efficiently. Due to these reasons, numerous clinical, industrial and experimental studies have been conducted. However, a clear comprehension of mutational effects is far from being complete. As a result, new high-throughput experimental and computational methods are highly needed to cover this unmet demand.

Among all human variants, rare missense changes make over 99% of the set of observed missense variants (with a Minor Allele Frequency, in short MAF, below 0.5%), and 90% are extremely rare (with MAF < 10^-6^)(9). This means that common variants can be more confidently annotated as truly benign and are therefore likely to offer higher labelling accuracy. Hence, the challenge primarily lies in accurately distinguishing between rare pathogenic variants and rare benign polymorphisms, which is crucial in assessing the pathogenicity of missense variations.

In recent years, multiple deep mutational scanning (DMS) experiments for various proteins were conducted by different groups. They demonstrated to experimentally assess the functional impact of nearly all possible missense variants for a target protein. However, these large-scale experiments are not easily implementable nor inexpensive in terms of time and costs. Therefore, highly efficient computational methods are in demand to predict the effects of non-synonymous mutations, and in particular missense mutations.

Many computational methods predicting mutational effects exploit information coming from multiple sequence alignments (MSA) and sequence covariation (10,11), and more recently deep learning models (12–19). Here, we are using information coming from three different biological scales: the evolutionary scale, the human population scale and the molecular scale. We exploit the evolutionary distance between natural sequences which encodes both epistatic information and conservation of amino acids positions (20), and protein structural information coming from available model structures produced by AlphaFold (21,22) to introduce ESCOTT, an epistatic and structural model of mutational effects. ESCOTT evolutionary- and structure-based predictions are coupled with information on allele frequencies collected in the Genome Aggregation Database (gnomAD) through an intuitive model privileging population-specific allele frequency. This results in a population-aware predictor of mutational effects, named PRESCOTT (Population-awaRe Epistatic and StruCtural mOdel of muTational effecTs). PRESCOTT handles common variants towards their labelling as benign mutations and assigns ESCOTT scores (no change) to rare variants towards their classification in benign and pathogenic. PRESCOTT relies on a very few parameters whose thresholds are computed on a small set of a few hundred proteins. No training on human clinical data over large quantities of rare pathogenic and rare benign variants is realized. Hence, PRESCOTT remains interpretable in each step of the computation process towards an understanding of the effect of a mutation.

## RESULTS

### ESCOTT models residues in sequences with evolutionary conservation and structural features

The genes observed across species today are products of extensive evolutionary processes that selected for functional molecules (**Figure 1A**). By modelling the constraints of these evolutionary processes on protein sequences, we can infer which mutations might be benign and which ones might be pathogenic, through an analysis that covers every possible substitution at every position (**Figure 1B**). To do so, ESCOTT exploits both the vast array of sequences and structural models available today. It models a protein sequence position by integrating two structural features with the evolutionary conservation of the position (23,24). The first structural feature focuses on the packing density of amino acids within the protein’s core. This aspect is crucial since mutations that change hydrophobic residues into hydrophilic residues, or those that modify amino acid size, can destabilize the protein, leading to a loss of function (25). This feature is quantified by the residue’s Circular Variance (CV), which measures the atomic density surrounding a residue, providing insight into its structural position (24,26). Lower CV values indicate a protruding residue while higher CV values indicate a buried residue. The second structural feature concerns changes of surface residues, which could impact the protein’s interaction with other molecules, potentially resulting in functional loss (27–29). This is assessed by the physico-chemical (PC) properties of the residues at an interface, where PC is a metric indicating the propensity of each amino acid to be located at a protein interface (30). The notion of evolutionary conservation, termed T_JET_ and adopted by ESCOTT from (23,24), is illustrated in **Figure 1C**. Here, the amino acids S, E, P, and V occur at position j in five maximal subtrees, each maintaining the same amino acid across all species within the subtree. This local residue conservation within the tree, captured by the size of the maximal subtrees conserving a residue at that position, is used in T_JET_ to model the conservation of a position j as a function of the sizes of the associated maximal subtrees. Thus, position j is deemed to be more conserved than position k if the residues in j are associated with larger subtrees than those in k. Therefore, for each residue, ESCOTT combines T_JET_ with the PC and CV metrics to identify (with higher numerical scores) those residues that are conserved or/and located either at the protein surface or within its core (**Figures 1D** and **1E**) as being more susceptible to bear pathogenic mutations. Additionally, for regions of the sequence forming coiled structures, ESCOTT exclusively models their evolutionary conservation using T_JET_.

**Figure 1.**
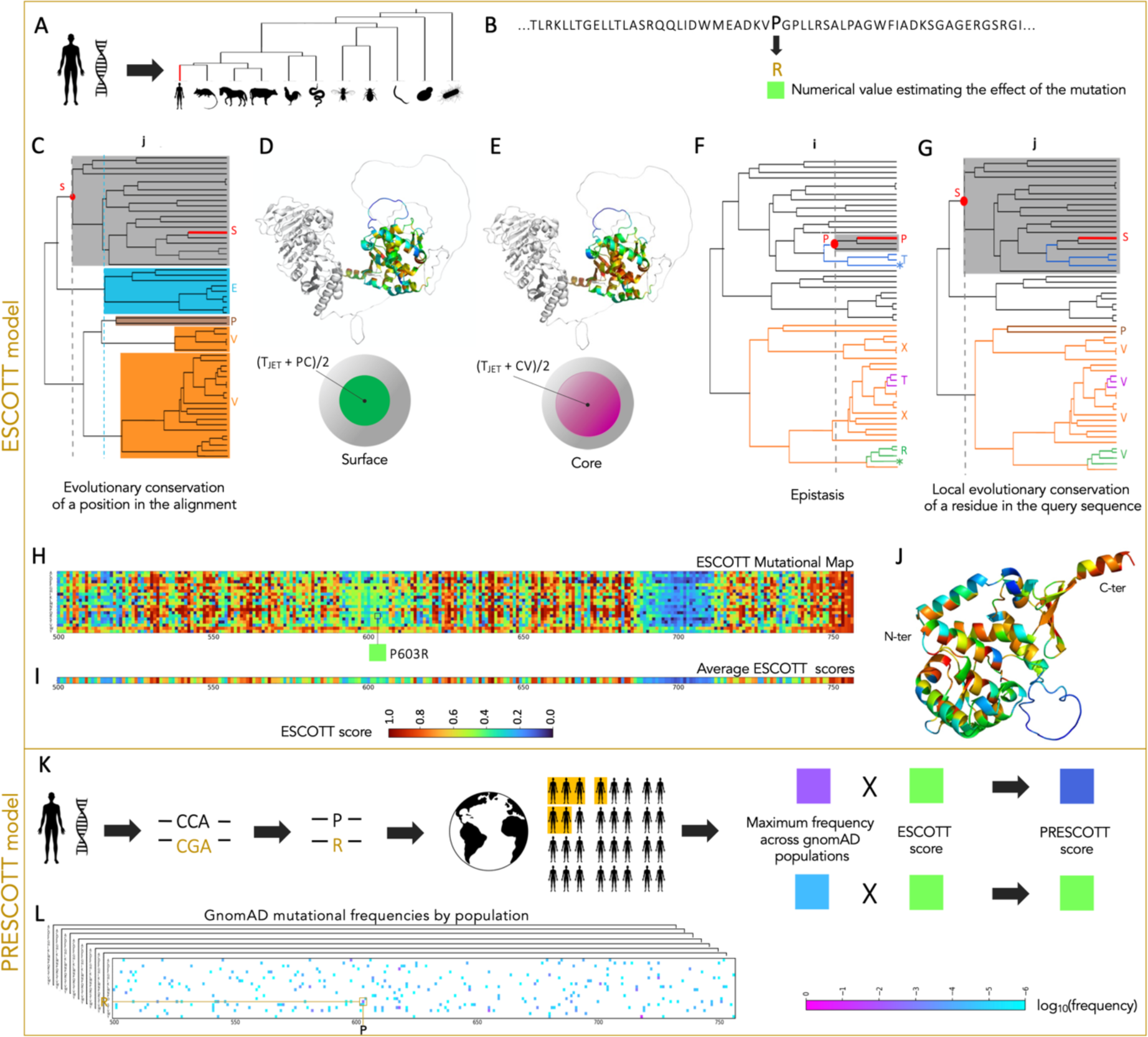
Conceptual building blocks in ESCOTT and PRESCOTT. The ESCOTT model is described in panels A-J and PRESCOTT in K-L. **A.** Given a protein sequence, for example in humans, ESCOTT examines its homologs and reconstructs a phylogenetic tree of sequences on which it evaluates the positions of the mutation in other species in the tree. The position of the query sequence in the tree is indicated by the red edge. **B.** The effect of a mutation, such as P-to-R at position i, will be predicted based on sequences in the tree (panels CFG) and on the model structure for the query sequence, if available (panels D and E). **C.** ESCOTT estimates an evolutionary conservation term (T_JET_): for position j in the query sequence, occupied by the amino acid S, ESCOTT considers the level in the tree (dashed line - grey subtree) where the amino acid at that position appeared and remained conserved thereafter. As in (Laine, Karami and Carbone 2019; (23), ESCOTT defines the evolutionary conservation of position j by looking at the height of maximal subtrees within the whole tree where the position is conserved, not necessarily with the same amino acid: here, there are five such maximal subtrees (colored grey, cyan, brown and orange). Two subtrees of maximum height (grey and cyan) are used to set the evolutionary conservation of the position. **D-E.** Positions 500-756 of the MHL1 human protein are colored in the structure with respect to the two terms used in ESCOTT model to evaluate residue positions by combining evolutionary conservation (T_JET_), physico-chemical properties (PC) and positions that belong to the structural core (CV). **F.** ESCOTT estimates an epistatic term that evaluates the effect of a mutation P-to-R with the minimal global amount of changes needed to “accept” R within species in the tree. The further the sequences accepting R in the tree (green) are from the reference one (red), the greater the mutational effect. **G.** ESCOTT compares sequence positions by favoring residues which are more conserved in the tree as shown for residue S at position j (panel E) vs residue P at position i (panel D). **H.** ESCOTT mutational map records mutational effects for all the 500-756 positions of the MHL1 human protein. Mutation P-to-R at position 603 is highlighted and the score reported with the corresponding color code. **I.** Averages of the scores by columns (across 19 mutations) are reported for positions 500-756 of MHL1. **J.** MLH1 structure colored with average scores in **I**. **K.** PRESCOTT combines ESCOTT scores and allele frequency in human populations, depending on whether allele frequency is higher than a fixed threshold. **L.** Allele frequencies are computed for the eight populations in gnomAD and a PopMax model employed. Each missense mutation is analysed independently with respect to the eight populations.

### ESCOTT traces structural information in the course of evolution to predict mutational effects in proteins

In our analysis, evolutionary processes satisfy two basic hypotheses. First, mutations appearing very rarely in nature are likely deleterious or, in other words, the more a residue is conserved across species, the more sensitive it is to mutations. Second, residues in a protein depend on other residues, because of structural and functional constraints, hence the effect of a given mutation depends on the presence or absence of other mutations. ESCOTT uses these two hypotheses to model the effect of a mutation, say P-to-R, at position i of the query sequence q, by analysing the distribution of aligned sequences organised on a distance tree with two main evolutionary measures (20) capturing 1. the minimal distance in the tree from q to a sequence which accepts R at position i, and 2. the different level of conservation for residues within the query sequence. These two measures are illustrated in **Figures 1F and 1G**, respectively. **Figure 1F** illustrates, for a position i occupied by residue P in q, the distance within the tree to the closest sequence, say s, displaying R at position i. The intuition is that if s is located far from q in the tree, the P-to-R mutation should require a large number of mutations in q for R to be “accepted” and therefore P-to-R should probably be deleterious. **Figure 1G** shows that when an amino acid S at position j in q is more evolutionarily conserved than the amino acid P at position i in q, the effect of a mutation for S should be greater than for P. The intuition is that mutations in positions that are locally conserved in large maximal subtrees should have more impact than mutations in positions associated with smaller maximal subtrees.

ESCOTT integrates its unique residue representation, which combines together the T_JET_, PC and CV values for the residue, into this global epistatic model of mutational effects describing mutational changes in the evolution of natural sequences. By doing this, ESCOTT evaluates the significance of each residue within the distance tree, with a particular focus on residues that are likely part of the protein’s surface or core structure. Notice that a residue does not need to be conserved to be considered significant. Indeed, ESCOTT considers a holistic set of factors for each residue, including its T_JET_, PC, and CV contributions, as well as the mutational context. This comprehensive approach allows ESCOTT to provide users with detailed insights into the effects of each mutation and to ensure transparency and depth in the analysis towards the final predictive score. This level of detail empowers users to gain a thorough understanding of the potential mutational impacts.

### PRESCOTT integrates population-specific allele frequencies to filter common variants

Genomic information from unrelated individuals in human populations across the world provides a complementary source of data for assessing the effect of a mutation. Because of its widespread use in clinical diagnosis, this data has been collected, through a community effort across countries, in the Genome Aggregation Database (gnomAD) (31). gnomAD contains mutation frequencies from unrelated individuals as part of human population genetics studies. It contains data from eight different populations: African/African American, Admixed American, Ashkenazi Jewish, East Asian, European (Finnish), European (non-Finnish), South Asian, and Middle Eastern. This information, when available, is generally used by experts to assess the significance of missense mutations. However, it remains intrinsically limited because allele frequencies depend on the number of individuals sequenced and other factors, which means that simply knowing allele frequencies is an uncertain basis for making decisions.

PRESCOTT combines ESCOTT predictions with population allele frequencies, providing a doctor with an integrated score for missense mutations (**Figure 1K**). To do this, PRESCOTT follows the common intuition that the higher the number of individuals carrying the mutation present in gnomAD, the lower the probability that the mutation is pathogenic. If a mutation is too frequent in relation to an overall allele frequency threshold (see Methods), PRESCOTT adjusts the ESCOTT score by decreasing it proportionally to the allele frequency recorded in one of the eight gnomAD populations, thus reinforcing the benign effect of the mutation. After evaluating a mutation on each population, PRESCOTT selects the popmax allele frequency, defined as the maximum allele frequency in the eight continental populations, as recommended in (32) (**Figure 1L**). As a general rule, if a variant is common in one population, it can be assumed to be benign across all populations.

The PRESCOTT model operates using two primary parameters, fine-tuned using sequenced data from gnomAD for a set of 500 human proteins. These parameters include a scaling coefficient, “c,” fixed at 1, and a frequency cutoff, “Fc,” established at 0.0001 (details in Methods section). The model doesn’t apply the frequency cutoff in a straightforward manner to filter variants. This is especially true for variants with a MAF less than 0.005 (0.5%), typically classified as rare. PRESCOTT evaluates all mutations with a frequency value f greater than 0.0001 (as per the testing condition “f>Fc” outlined in Methods), including those between 0.005 and 0.0001. The model adjusts the ESCOTT score of these mutations by the factor “[Fc - f]/Fc.” Therefore, a mutation with a high ESCOTT value, indicating potential pathogenicity, is less likely to have its clinical assessment altered due to its low frequency. Conversely, for mutations that have lower ESCOTT scores, indicative of benign effects, incorporating allele frequency data reinforces the prediction and leads to the reclassification of certain Variants of Uncertain Significance (VUS) as benign.

Figure 2 illustrates the intermediate steps following ESCOTT score matrix reconstruction for the MutL protein Homolog 1 encoded by the *MLH1* gene in the human genome, which is involved in DNA repair. Diseases associated with *MLH1* include Mismatch Repair Cancer Syndrome 1 and Lynch Syndrome 2. This protein is of high medical interest and allele frequencies of its missense mutations are included in gnomAD. It should be noted that the ESCOTT score matrix describes the impact for all mutations (Figure 2A), whereas genetic disorders relate only to identified missense mutations, which are one nucleotide away from the parental DNA sequence. This is a much smaller subset of mutations (compare with Figure 1L). Figure 2E illustrates how the use of allele frequencies from gnomAD (Figure 2C) is instrumental in converting an ESCOTT score (Figure 2B) into a PRESCOTT score (Figure 2D). It is crucial to understand that the PRESCOTT model selectively employs gnomAD allele frequency data to modify only those ESCOTT scores associated with sufficiently high allele frequencies, as detailed in the Methods section. Scores corresponding to lower frequencies are not adjusted in this process (Figure 2C, circled mutations). A key aspect of PRESCOTT’s methodology is its independence from any ClinVar classification that may be present in gnomAD, which typically categorizes variants as pathogenic, benign, likely pathogenic, or likely benign.

**Figure 2.**
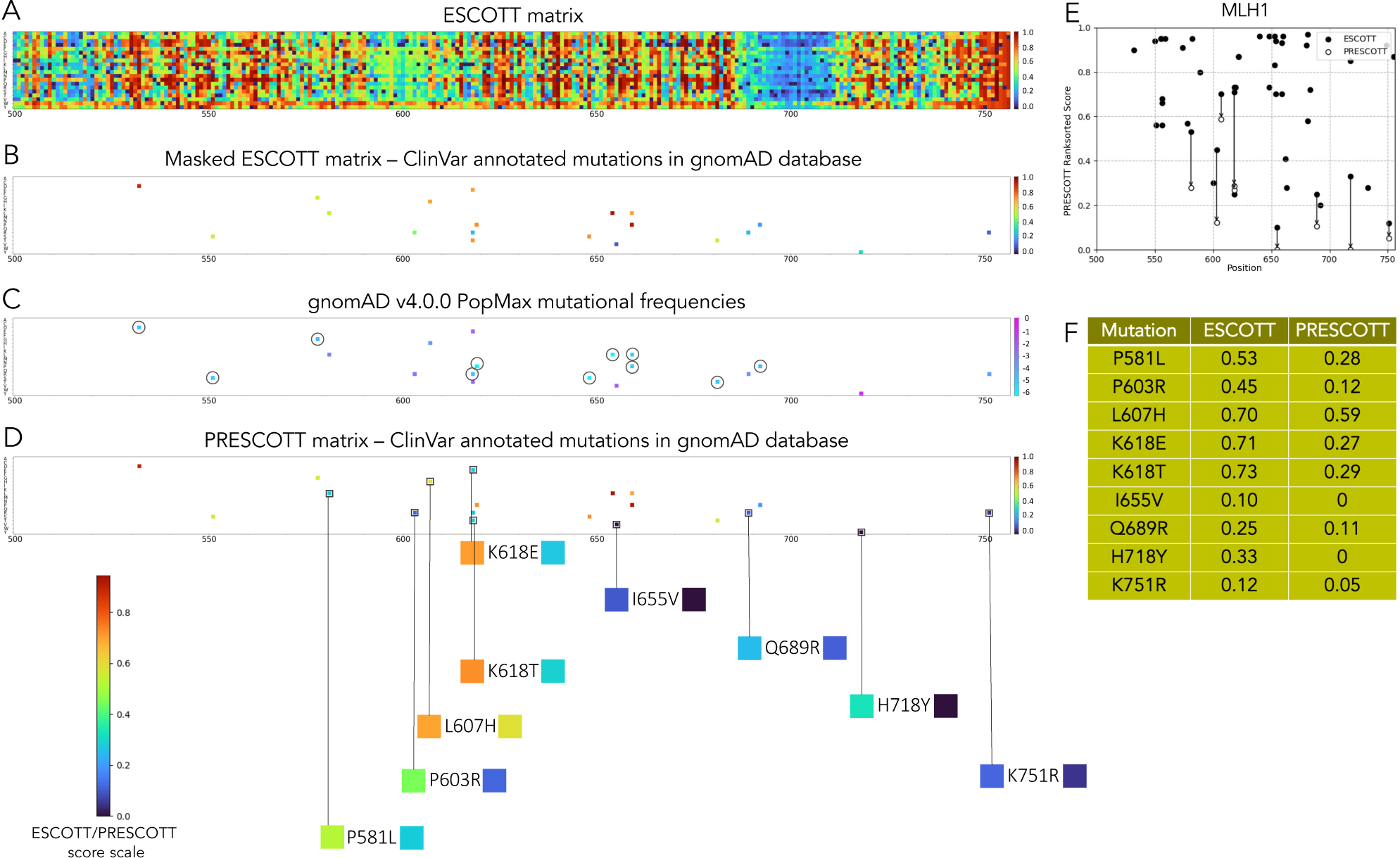
Analysis of the human MLH1 protein with PRESCOTT. We report the analysis for the 500-756 region of the MLH1 protein. For this region, we consider 20 mutations, that is all those gnomAD mutations that are labelled as benign/likely benign and pathogenic/likely pathogenic in ClinVar. **A.** Generation of the ESCOTT matrix defined by all possible mutations going from any amino acid position in the original sequence to any other amino acid among 20. **B.** ESCOTT matrix in **A** is masked to highlight only the scores for the 20 pathogenic or benign mutations in gnomAD. **C.** gnomAD v.4.0.0 matrix describing the maximum frequency for the 20 mutations in gnomAD across the eight gnomAD populations. Circled mutations have very low allele frequency. **D.** PRESCOTT matrix describing the scores for the 9 gnomAD mutations differing from ESCOTT scores. The change between ESCOTT (left colored square) and PRESCOTT (right colored square) scores is visualised by the colors below the matrix. **E.** Plot of ESCOTT scores (black circles) for 48 mutations in the region of the MLH1 protein covering positions 500-756. These mutations are described in ClinVar as benign/likely benign (16, blue) and pathogenic/likely pathogenic (32, red); note that only 45 points are visible due to 3 positions with double mutation (L555P, L555R both Likely pathogenic with ESCOTT score 0.88; L653P, L653R both Likely pathogenic, 0.87; R659L, R659P both Pathogenic, 0.75). These include the 20 mutations present in gnomAD. PRESCOTT scores (white circles) might lower some ESCOTT scores as indicated by the arrows. Compare to **Figure 7BC**. **F.** ESCOTT and PRESCOTT scores for 9 gnomAD mutations out of 20 show different values due to a high allele frequency estimation which is considered in PRESCOTT.

### A large-scale assessment of ESCOTT highlights an accurate identification of sensitive regions in proteins

ESCOTT produces full length sequence evaluations for all possible mutations of a protein sequence. It can be applied to large proteins, intrinsically disordered regions, splicing variants, mutated sequences, artificially generated sequences. The method is sensitive to the long-range correlations existing for amino acid positions, making it possible to detect differences in the mutational landscape of close sequences. We tested the performance of ESCOTT on 32 human protein DMS experiments available in the ProteinGym database, and its generality across species on all the 87 DMS experiments.

We compared ESCOTT with the 3 state-of-the-art methods EVE_ensemble (13), ESM1b (18), and AlphaMissense (19) by computing the average Spearman correlation between their predictions and the 32

DMS experiments on the human proteins of the reference ProteinGym dataset (12) (Figure 3 and **Table 1**). AlphaMissense has the same level of accuracy as ESCOTT with an average Spearman correlation coefficient of 0.46 calculated on 218,301 point mutations (**Table 1**). It should be noted that AlphaMissense has been fine-tuned on human and primate variant population frequency data and the confidence calibrated on known disease variants. In contrast, ESCOTT only relies on sequence and structure information. An extended comparison of ESCOTT with 37 existing methods, including GEMME and iGEMME, is reported in Figure 3B.

**Table 1.** Comparison of ESCOTT with EVE (ensemble), ESM1b and AlphaMissense on human proteins in the ProteinGym dataset. Average Spearman correlation coefficients are reported for all, single and multiple mutation experiments on the 32 human proteins in the ProteinGym dataset. “Average” refers to Spearman correlation computed on the average vector associated to a matrix. For each subset, the best result is in bold and the second best is underlined.

| Mutational classes | ESCOTT | EVE (ensemble) | ESM1b | AlphaMissense |
| --- | --- | --- | --- | --- |
| Overall | **0.460** | 0.419 | 0.405 | **0.460** |
| Single | <u>0.451</u> | 0.410 | 0.403 | **0.456** |
| Multiple | **0.524** | 0.482 | 0.420 | <u>0.493</u> |
| Average (single) | **0.573** | 0.520 | 0.515 | <u>0.522</u> |

**Figure 3.**
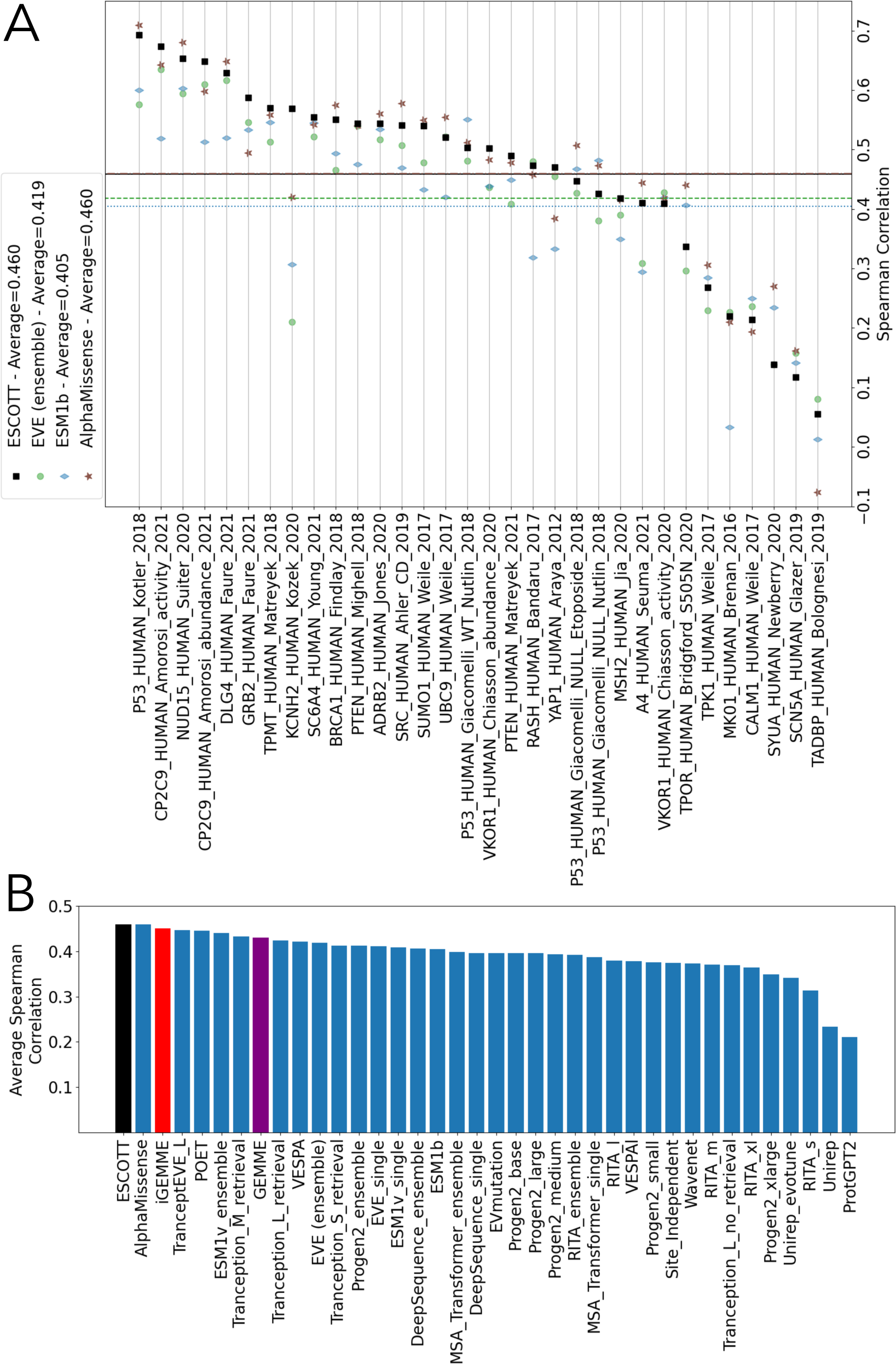
Comparison of ESCOTT, EVE (ensemble), ESM1b and AlphaMissense on 32 DMS experiments for human proteins. **A.** ESCOTT (black squares), EVE (ensemble, green circle), ESM1b (blue diamond) and AlphaMissense (brown stars). Dashed lines indicate average Spearman correlation coefficients of the predictive methods with experimental data from the 32 human DMS experiments in the ProteinGym dataset: solid black line for ESCOTT, dashed dotted brown line for AlphaMissense, dashed green line for EVE (ensemble), dotted blue line for ESM1b. **B.** Comparison of ESCOTT (black) with existing methods. The list of methods includes EVE (ensemble), AlphaMissense, iGEMME (red), GEMME (purple), and ESM1b. Data for all other methods was obtained from https://www.proteingym.org/substitutions-dms-level, except for AlphaMissense and PoET (ensemble) which are reported in (19) and (75) respectively. SPAST_Human (Q9UBP0) - Spastin

To gain a deeper understanding of the matrices produced by ESCOTT, EVE_ensemble, ESM1b, and AlphaMissense, we have focused on those consecutive positions in a matrix for which most of the amino acid mutations display high scores. These high value scores are indicators of pathogenicity. An example of “sensitive” regions in a protein, that is regions detected by the methods as likely to host pathogenic mutations, is shown in Figure 4, for the human Spastin protein (encoded in the *SPAST* gene), previously used in (18) to compare ESM1b to EVE. The score matrices for the four computational approaches applied to the human Spastin are displayed in Figure 4A-D. As remarked in (18), ESM1b highlights one main sensitive region corresponding to the MIT domain (120–195) (Figure 4D). ESCOTT and AlphaMissense detect several other regions of interest, highlighted in the UniProt and InterPro databases (Figure 4AB). These are the MIT, ATPase domain (343–506), AAA+lid (534–571) and VPS4 (578–612) domains and three other regions, required for interaction with ATL1 (1–80), for interaction with SSNA1 and microtubules (50–87), and the ATP-binding motif (382–389), along the entire length of the protein. The EVE analysis does not cover approximately half of the total length of the protein, but over the region provided, it highlights known biological signals (Figure 4C). Interestingly, the N-terminal region of the Spastin protein is marked by relatively high uncertainty by AlphaFold (Figure 4E) whereas both ESCOTT and AlphaMissense can recognise insightful biological signals there. In general, AlphaFold appears to reconstruct a structural model with greater confidence for regions of the protein that are also predicted with higher ESCOTT values.

**Figure 4.**
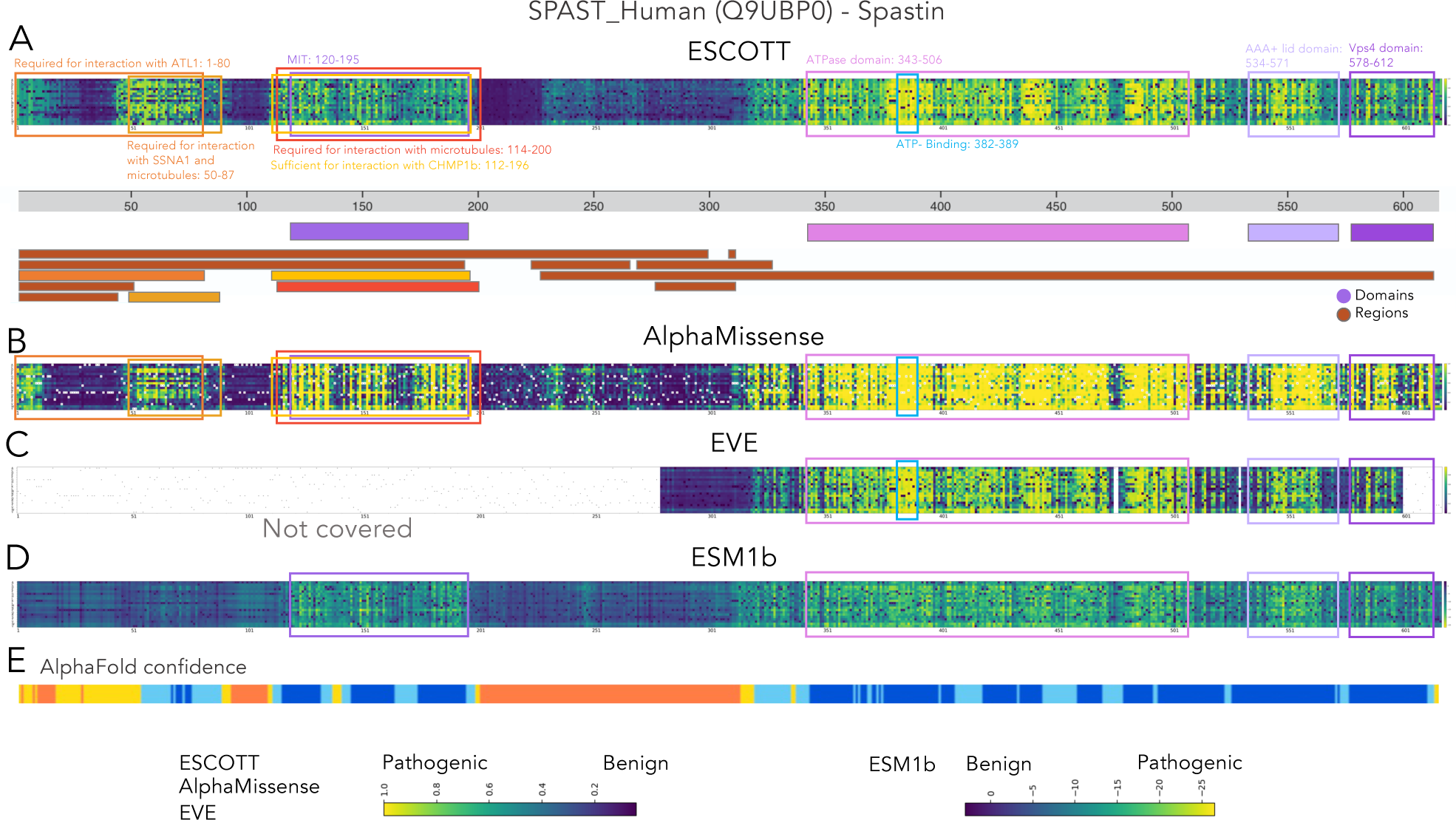
ESCOTT highlights functionally relevant regions of proteins. **A.** The human *SPAST* gene, encoding the spastin protein. ESCOTT evaluation of the full length spastin protein. The matrix reports ranksorted ESCOTT scores in a colorscale going from dark blue (0, no effect) to yellow (1, highest effect). Below the ESCOTT matrix, known domains (purple color scale) and regions (brown color scale) of interest for the protein are reported from the UniProtKB database (www.uniprot.org/uniprotkb/Q9UBP0/feature-viewer) and the InterPro database (www.ebi.ac.uk/interpro/protein/UniProt/Q9UBP0/), data download in August 2023. **B.** AlphaMissense analysis. **C.** EVE analysis of spastin. The four white regions have not been calculated by EVE. They correspond to approximately half of the positions. **D.** ESM1b analysis, as reported in (18). **E.** AlphaFold confidence (from InterPro) of positions in the spastin‘s structural model. Confidence is represented by a color scale from dark blue (very high) to red (very low).

It should be noted that sensitive regions identified in the ESCOTT matrix are not a direct consequence of the integration of structural information into the model. Indeed, the detection of sensitive regions for the SPASTIN protein is also present in the iGEMME matrix, which is based solely on evolutionary information, as shown in **Figure S1**. However, the use of structural information improves ESCOTT scores. In addition, the ESCOTT matrix based on ranksorted scores (see Methods) increases the intensity of the biological signal. Finally, it should be noted that a number of other regions along the protein, such as 437-447 and 470-487 in the ATPase domain, are highlighted by ESCOTT and EVE as being sensitive to mutations, but that no functional interpretation of these regions is known at present.

To obtain a quantitative assessment of the “sensitive regions” in a protein, that is those regions which are most sensitive to mutations, we computed the average of the scores per column in the four matrices, thus reducing the information in the matrix to a vector of average values. Then, we computed the average Spearman’s correlation coefficient of the 28 DMS matrices from single-mutation experiments (Figure 5A). For almost all proteins and all methods, correlation values increased sharply compared to the correlations computed on the full DMS matrices (Figure 3A). This means that all methods are able to identify the most sensitive regions for a protein. ESCOTT obtains an overall average across the 28 experiments of 0.573. This performance surpasses that of AlphaMissense, which scored 0.522, EVE with 0.520, and ESM1b at 0.515. These average scores can be visually represented on protein structures, aiding in the identification of sensitive regions. For instance, Figure 5B illustrates this with the Thiopurine S-methyltransferase protein (TPMT), where sensitive areas are highlighted in yellow tones. This three-dimensional plotting is instrumental to users in further analyzing proteins, particularly in identifying binding pockets, interaction sites, and functionally relevant residues. In the case of TPMT, both ESCOTT and AlphaMissense exhibit consistency with experimental values. They show comparable correlations in their experimental predictions and consistently identify the same sensitive areas within the protein’s structure.

**Figure 5.**
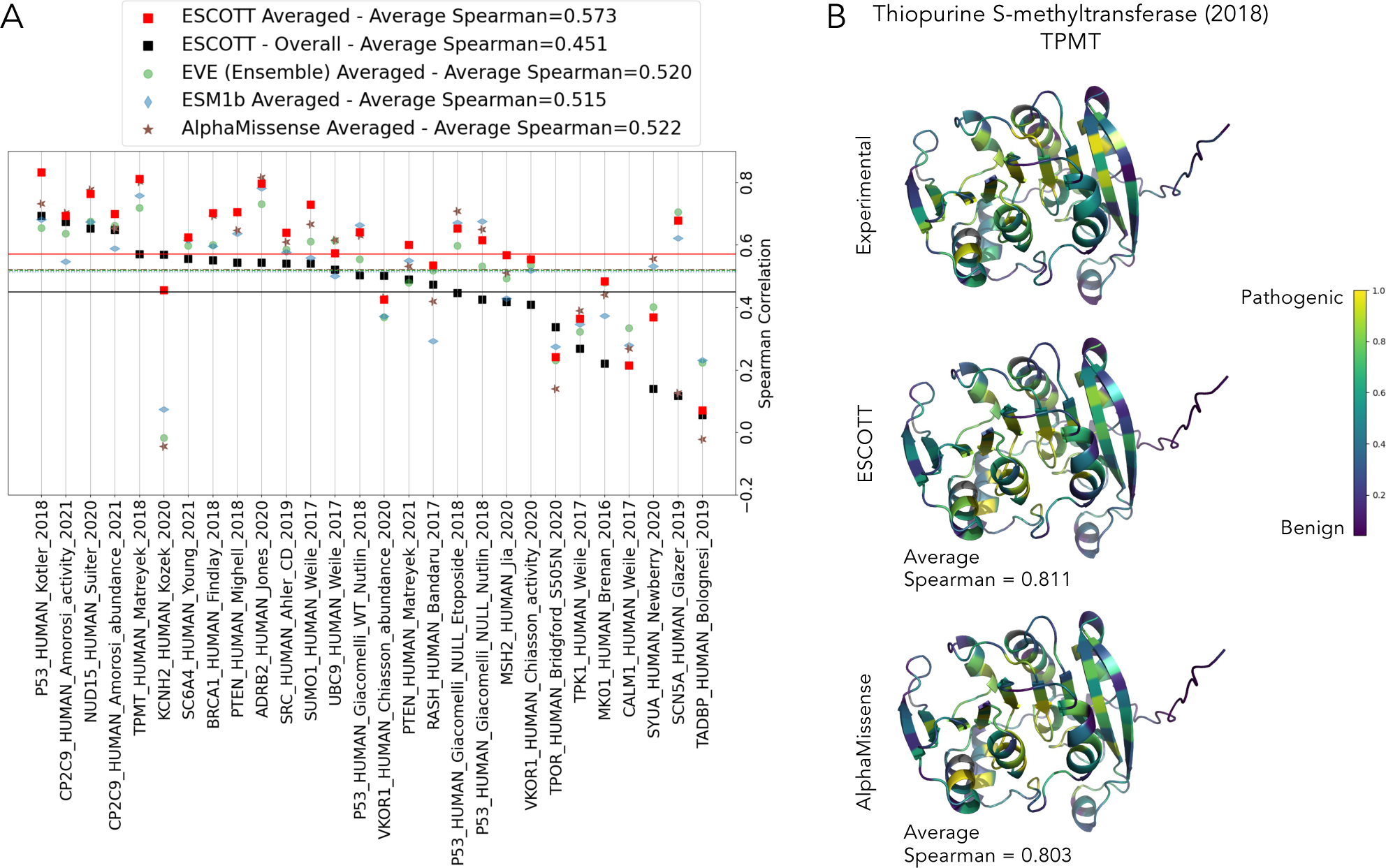
ESCOTT highlights functionally relevant regions of proteins. A. Spearman correlation coefficients between positional averages of the 28 single mutation DMS matrices and predicted matrices in ProteinGym. Black squares correspond to the values reported in Figure 2 for ESCOTT. B. Averaged scores from experiments, ESCOTT and AlphaMissense are projected onto the human Thiopurine methyltransferase (TPMT_HUMAN in A) enzyme. A viridis color scale was used for low (blue) versus high (yellow) average scores projected onto the protein structure. Grey residues (in positions 95, 96, 117, 172) are associated to absence of experimental data. Protein visualizations were realized with

ESCOTT performance has also been evaluated on the entire ProteinGym dataset of 87 DMS experiments. A comparative account of the four methods on each experiment is shown in **Figure S2A**, and a more global comparison with 37 state-of-the-art methods (10,12–17,33–40), is reported in **Figure S2B**. AlphaMissense outperforms all methods. Yet it is important to note that its advantage partly stems from being explicitly fine-tuned to distinguish between pathogenic and benign effects. In contrast, ESCOTT, along with the other comparative methods, does not utilize classified data in its modeling process. The incorporation of structural information into the ESCOTT model marks a significant improvement over iGEMME and GEMME. These two models are ranked just below POET and TranceptEVE_L. Thanks to this structural integration, ESCOTT secures the position of second best after AlphaMissense. It achieves a correlation coefficient of 0.492 over the entire ProteinGym dataset, closely following AlphaMissense, which scores 0.525.

A comparative account of the four methods on the average Spearman correlation coefficient for the 76 single point DMS experiments across species is reported in **Figure S3**. Consistently with what we already observed for human proteins, all methods improve the prediction of sensitive regions compared to the base line fixed on full matrix correlations.

### Some global observations lead to an improved interpretation of ESCOTT scores

Are sensitive positions located in three-dimensional clusters? An answer to this question would be particularly useful in interpreting protein mutations affecting specific positions or spatial regions in a protein. A visual inspection of the human Thiopurine methyltransferase (TPMT) (Figure 5B) shows contiguous three-dimensional regions within the protein structure that are sensitive to mutations, suggesting that a protein displays high sensitivity in specific three-dimensional regions and that these regions can be predicted with high accuracy (ESCOTT average Spearman is 0.811 and AlphaMissense is 0.803 on TPMT). In conclusion, ESCOTT scores over amino acid positions are highly informative about the sensitivity of the protein to mutations and allow the identification of sensitive regions along the sequence but also within the structure.

Can ESCOTT better predict phenotypes induced by mutations located in preferred structural elements, such as helices, beta strands, or coils? We evaluated ESCOTT’s predictions of mutations occurring in different structural elements and compared them with experimental data collected at the same locations. The average Spearman correlation increases from 0.489 to 0.497 for mutations in structured regions (specifically at positions annotated as secondary structural elements H, E, G, B, I, S, T in DSSP notation). In contrast, for coiled regions (longer than 5aa; see Materials and Methods; **Figure S4**), the average Spearman correlation drops to 0.450. In conclusion, our analysis suggests that ESCOTT better predicts the effect of mutations located in structured regions.

Are mutations towards specific amino acids better predicted, regardless of the starting amino acid? To this end, we computed the Spearman correlations for mutations toward a specific amino acid in all single mutational experiments of the ProteinGym dataset. **Figure S5** shows that ESCOTT predicts mutations toward hydrophilic residues (D, R, E, K, N, Q) particularly well, with the best prediction for positively and negatively charged amino acids D, R, E, K. On the other hand, Spearman’s average reaches the lowest values for hydrophobic residues (I, V, L, Y, F, W, M, C), reducing to 0.330 for C. The predictions of mutations toward neutral residues (G, S, P, T, H, A) (where P is poorly hydrophilic) are consistently better than those for hydrophobic residues. In conclusion, prediction of mutational effects for specific amino acid changes improves accuracy.

### A large-scale assessment of PRESCOTT that uses population-specific counting of allele variants

A major interest in clinical research is to distinguish, in an individual’s genome, a rare variant that causes disease from the millions of common and largely benign variants found in every human genome. Today, the gnomAD v4.0 database contains variant data on more than 807,162 individuals. However, a very small part of missense variants are currently observed in gnomAD (estimated to ∼12% in 2022 (32)), indicating that much larger numbers of individuals will need to be sequenced before the full spectrum of tolerated variants can be discovered (32).

The majority of rare variants are not pathogenic, and current reference population databases have not yet reached saturation for most types of variation (41). Furthermore, genetic studies often disproportionately represent European populations, leading to an underrepresentation of other communities. This underrepresentation causes individuals from these communities to have a higher number of rare variants with uncertain significance (VUS). Additionally, datasets may include individuals with Mendelian diseases, and despite stringent quality control, sequencing or annotation errors can occur. Given these factors, there is a pressing need for computational methods that are not only interpretable but also capable of providing detailed explanations for mutations suspected to be pathogenic and for regions sensitive to mutations. PRESCOTT has been developed to meet these challenges. It focuses on processing common variants towards categorizing them as benign and minimizes the number of rare variants, specifically evaluating those with allele frequencies between 0.005 and 0.0001. This is achieved by integrating basic allele frequency data, a practice well-established in clinical settings, with the ESCOTT model’s computational predictions that are based on sequence and structure.

We evaluated PRESCOTT’s effectiveness. This evaluation involved assessing PRESCOTT’s capability to reliably assign higher scores to pathogenic mutations compared to benign ones. Utilizing a comprehensive dataset of 1,883 proteins, as detailed in **Table 2** in Methods, with 7,954 pathogenic and 9,276 benign mutations from ClinVar, PRESCOTT demonstrated its proficiency without prior knowledge of their ClinVar categorization. It achieved an impressive AUC of 0.95. This performance was benchmarked against other models: AlphaMissense (0.91), EVE (0.87), ESM1b (0.87), and ESCOTT (0.88), as illustrated in Figure 6A. The superior performance of PRESCOTT underscores the critical role of allele frequency data in refining ESCOTT’s scoring system. Moreover, the analysis of the two distinguished ClinVar sets of pathogenic and benign mutations reported in Figure 6B, clearly show that PRESCOTT model engraves on the ESCOTT distribution of scores for benign mutations (see Methods for the algorithm, the parameters, and the evaluation of the parameters’ thresholds on an independent dataset of proteins). Notably, AlphaMissense also predominantly assigns lower scores to benign mutations. However, it differs from PRESCOTT as it assigns a considerable number of low scores to pathogenic mutations as well. Models like EVE and ESM1b, which do not incorporate allele frequency information, predictably exhibit lower performance, similar to ESCOTT. Application of the PRESCOTTscore-low-scores algorithm to the EVE model resulted in a marked improvement elevating EVE’s AUC to 0.95. Additionally, we conducted an analysis using ClinVar’s starred subsets, which reflect a higher confidence level due to consensus from multiple laboratories. This analysis, depicted in **Figure S6**, aligns well with our primary findings and offers valuable insights into data quality criteria.

**Figure 6.**
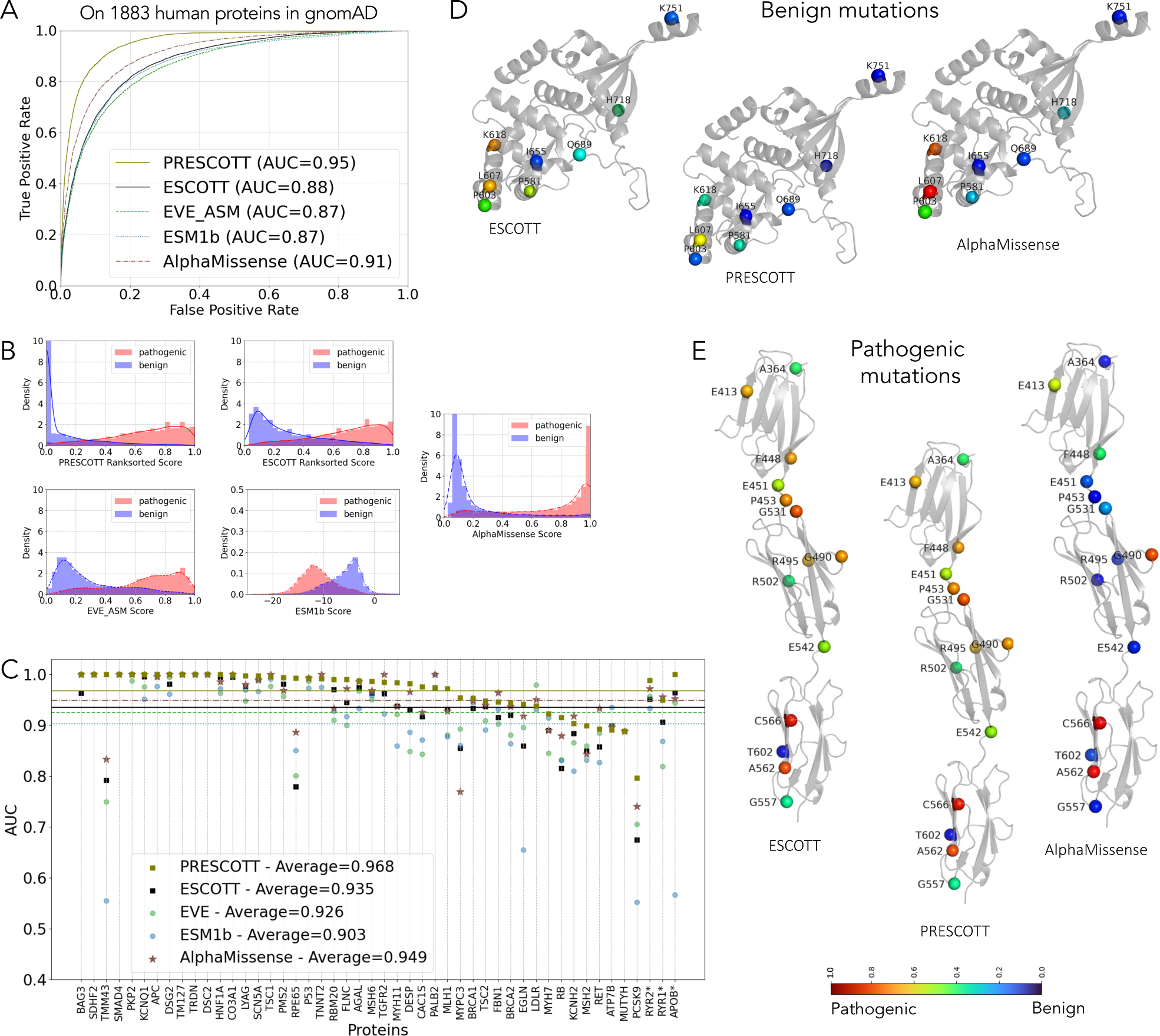
Comparison of PRESCOTT, ESCOTT, EVE, ESM1b and AlphaMissense on human proteins in gnomAD and ACMG datasets. **A.** Analysis of 1,883 human proteins whose mutations collected in gnomAD v4.0.0 are labelled as pathogenic or benign in ClinVar. AUC curves for PRESCOTT, ESCOTT, EVE, ESM1b and AlphaMissense are superimposed. **B.** PRESCOTT, ESCOTT, EVE, ESM1b and AlphaMissense distributions of scores for the 17,230 mutations of the 1,883 human proteins. The plots split scores in two disjoint subsets, for mutations labelled as benign (blue) or pathogenic (red) by ClinVar. **C.** Global comparative analysis based on AUC scores for the 48 human proteins in the ACMG v3.1 dataset, for ESCOTT, PRESCOTT, EVE, ESM1b and AlphaMissense. Note that there is a structural model for 45 of the 48 proteins. The three remaining proteins, with a starred name in the figure, RYR2=5050 aa, RYR1=5122 aa and APOB=4640 aa, have been evaluated with iGEMME. Note that all 48 proteins are represented by both benign and pathogenic mutations in ClinVar, and that both types of mutation are required to calculate the AUC and make the comparison. All proteins **present at least 3 benign mutations in ClinVar.** Compare with **Figure S7** reporting the same analysis on 64 human proteins with at least 1 benign mutation described in ClinVar. **D.** ESCOTT, PRESCOTT and AlphaMissense scores of 8 mutations are plot onto the MLH1 structure. These 8 mutations are labelled as benign/likely benign in ClinVar and are listed in **Figure 2G**. Due to multiple mutations at position 618, only K618T is plot. ESCOTT and PRESCOTT scores are reported in **Figure 2G**. Color scale is as in **Figure 2**. **E.** ESCOTT, PRESCOTT and AlphaMissense scores of 14 mutations annotated as pathogenic/likely pathogenic in ClinVar are plotted onto the MYPC3 structure. ESCOTT and PRESCOTT displays show identical scores.

### PRESCOTT on the ACMG dataset

PRESCOTT has been assessed on the 64 actionable human genes from the American College of Medical Genetics and Genomics dataset (ACMG, version 3.1) (42). This dataset contains 3,790 pathogenic mutations and 1,353 benign ones, which were all manually curated and certified by multiple sources in ClinVar. After generating model structures with Colabfold (43–46) and Alphafold (47) (see Methods), we ran PRESCOTT and compared it to EVE performance as reported in (13), ESMB1b and AlphaMissense. Figure 6C shows that on all ACMG proteins, PRESCOTT achieves an average AUC of 0.968, consistent with its performance on the 1,883 human proteins labelled in gnomAD (Figure 6A). (See also **Figure S7**.) Figures 6D and **6E** showcase the application of PRESCOTT on two specific proteins. Figure 6D focuses on a set of ClinVar-classified benign variants of the MLH1 protein (see Figure 2). Here, we observe notable improvements in the scores assigned by PRESCOTT compared to those by ESCOTT. This is particularly evident in the case of mutations K618 and L607, predicted as pathogenic by AlphaMissense, which are re-evaluated effectively in PRESCOTT thanks to the incorporation of allele frequency data. Conversely, Figure 6E presents an analysis of 14 ClinVar-identified pathogenic variants in the MYPC3 protein. In this instance, both ESCOTT and PRESCOTT assign identical scores to all mutations, attributable to the low allele frequencies that are not factored in by PRESCOTTscore-low-scores algorithm. Notably, some variants like A364, R502, G557, and T602 receive low scores from ESCOTT, indicating a failure to detect their pathogenicity. In comparison, AlphaMissense consistently assigns very low scores to almost all these variants. This results in a lower AUC of 0.769 for the entire MYPC3 protein, as opposed to the significantly higher AUC of 0.954 achieved by PRESCOTT (Figure 6C).

### ESCOTT and PRESCOTT intermediate scores to interpret human variants

ESCOTT and PRESCOTT produce a detailed array of intermediate scores for each mutation, which sheds light on the importance of residues from multiple angles, including their evolutionary conservation, location in the structure (core or surface), and physico-chemical properties. Moreover, the differences between ESCOTT and PRESCOTT scores offer insights into the population-level variations among variants. Pathologists can leverage these four distinct scores to assess mutations’ potential pathogenicity or benignity, aiding in the decision-making process for necessary laboratory tests for loss- or gain-of-function. These intermediate scores play a crucial role in deciphering the mechanisms behind mutations, contributing significantly to the overall evaluation made by both ESCOTT and PRESCOTT.

### Classification of Human Variants as Benign, Pathogenic, or VUS by ESCOTT and PRESCOTT

Recognizing the necessity for automated classification of variants into benign, pathogenic, and VUS, we established two thresholds. These were derived from the analysis of 1,849 pathogenic and 2,523 benign mutations associated with the 500 human proteins in our training dataset. By examining the precision and recall curves based on their predicted PRESCOTT scores, we identified thresholds that yield 90% precision across these proteins (shown in Figure 7A). Mutations with scores below 0.28 are categorized as benign, while those above 0.42 are considered pathogenic. Scores falling in between these thresholds are indicative of VUS. In Figure 7B, we apply these thresholds to the MLH1 protein’s 500-756 region, as seen in Figure 2E. Notably, several variants shift from the pathogenic (red) to the benign category (blue) in PRESCOTT scores due to allele frequency data. Figure 7C color-codes these mutations according to their ClinVar classifications, revealing that the mutations with lowered PRESCOTT scores align with benign classifications in ClinVar. PRESCOTT’s automated labelling of mutations as benign, pathogenic, or VUS shows reasonable agreement with ClinVar classifications.

**Figure 7.**
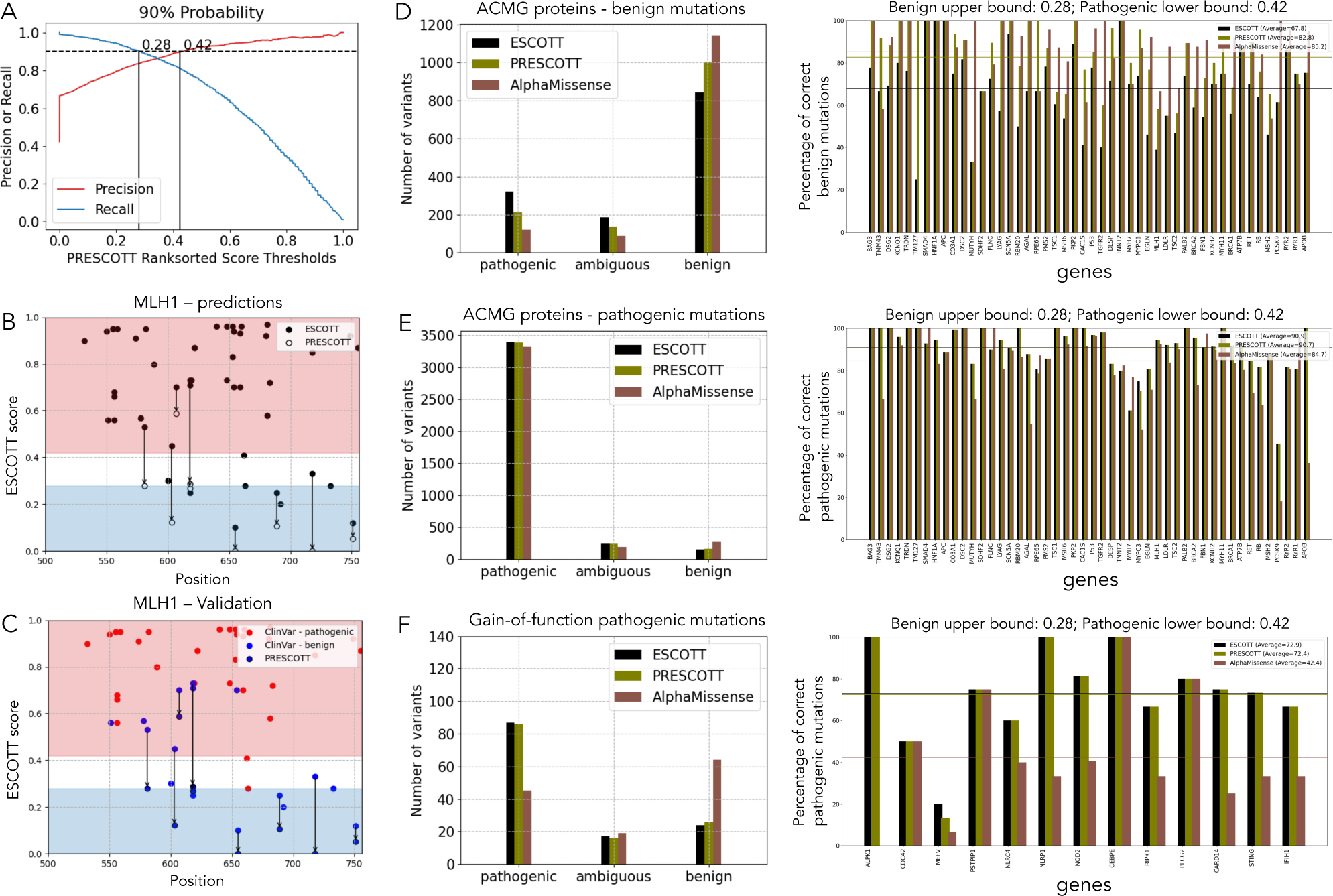
ESCOTT and PRESCOTT benign versus pathogenic mutations. A. Precision and Recall curves for the 500 human proteins used to calibrate the two thresholds in ESCOTT and PRESCOTT for the purpose of distinguishing between benign and pathogenic variants. B. Same as Figure 2E where a colored background indicates those mutations that are labelled as pathogenic (red), benign (blue), VUS (white) variants by ESCOTT/PRESCOTT. C. The 48 mutations considered in B are colored with ClinVar classification as benign/likely benign (16, blue) and pathogenic/likely pathogenic (32, red). Only 45 points are visible, as described in the legend of Figure 2E. D. The set of ACMG benign mutations. The plot on the left reports the exact count of mutations which are wrongly predicted as pathogenic (left), or VUS (centre) and correctly as benign (right) by each method. The plot on the right reports, for each protein in the dataset, the percentage of correctly predicted mutations. The horizontal lines indicate the average over all percentages obtained by each method. E. The set of ACMG pathogenic mutations is analysed as in D. F. The set of gain-of-function pathogenic mutations involved in autoinflammatory diseases is analysed as in D.

An extensive comparison of ESCOTT/PRESCOTT and AlphaMissense classifications is presented in Figures 7D and **7E**, focusing on the ACMG dataset of human proteins. For the 1,353 benign variants, AlphaMissense outperforms PRESCOTT, correctly identifying 1,144 variants compared to PRESCOTT’s 1,005, and incorrectly labeling 121 as pathogenic versus PRESCOTT’s 210. However, in the larger pool of 3,790 pathogenic mutations, ESCOTT’s performance is superior, accurately predicting 3,391 variants compared to AlphaMissense’s 3,315, and misclassifying fewer variants as benign (161 versus 276 by AlphaMissense). Intriguingly, ESCOTT’s accurate predictions for pathogenic variants are achieved without relying on allele frequency data. To further underscore this observation, we examined a set of proteins known for gain-of-function pathogenic variants, which are notoriously challenging to predict.

### PRESCOTT on gain-of-function

In our focus on gain-of-function mutations, we assembled a dataset featuring pathogenic missense mutations with gain-of-function, sourced from thirteen human genes associated with autoinflammatory diseases. These genes include *ALPK1, CDC42, MEFV, PSTPIP1, NLRC4, NLRP1, NOD2, CEBPE, RIPK1, PLCG2, CARD14, STING*, and *IFIH1*. In Figure 7F, we present a comparative analysis of these mutations, totaling 128, between ESCOTT/PRESCOTT and AlphaMissense. The results show that ESCOTT/PRESCOTT successfully identifies over 67% of these mutations as pathogenic, whereas AlphaMissense recognizes only about 35% as such (Figure 7F, left side). A similar pattern is observed when evaluating a subset of 102 pathogenic mutations using EVE and ESM1b. These models slightly outperform AlphaMissense, with ESCOTT/PRESCOTT correctly classifying 71%, AlphaMissense 37%, EVE 52%, and ESM1b 49% of the mutations. The complete list of the 128 mutations, along with the scores assigned by ESCOTT/PRESCOTT, AlphaMissense, ESM1b, and EVE, is available in the supplementary file “gain-of-function-mutations.csv”. Additionally, Figure 7F (right side) details the predictions of ESCOTT/PRESCOTT and AlphaMissense for each specific protein. Notably, for the ALPK1 protein, which is represented by just two mutations, these mutations are wrongly predicted by AlphaMissense. The analysis of the 15 pathogenic mutations in the *MEFV* protein is particularly intriguing, as these mutations pose significant challenges for both ESCOTT/PRESCOTT and AlphaMissense systems.

### The PRESCOTT Web Server and Its Comprehensive Human Protein Database

The PRESCOTT web server, along with its extensive human protein database of over 19,000 human proteins pre-analyzed using the ESCOTT model, is expected to be a highly valuable tool in both research and clinical settings. Central to PRESCOTT is its pathogenicity scoring system, designed to assess the disease-causing potential of mutations. Users are empowered to customize their analyses by merging these precomputed ESCOTT analyses with either the latest gnomAD dataset or their personally specified allele frequency data to generate PRESCOTT scores. Additionally, the PRESCOTT webserver’s capability to conduct ESCOTT model calculations on proteins from various species greatly extends its utility and scope. Notably, the adaptability of the PRESCOTT model to different species, including primates, highlights its wide-ranging applicability.

## DISCUSSION

Missense variants involve single nucleotide substitutions and make up the overwhelming majority of VUS, which are of clinical interest in diagnosing disease-causing genes/mutations. The impact of these mutations on proteins is complex, affecting protein interactions, structural stability, and functional gains or losses. The manifestation of a missense variant as pathogenic in one individual and benign in another can be attributed to differences in genetic background, environment, and diet. The challenge lies in distinguishing potentially pathogenic variants from the vast majority of benign variants.

PRESCOTT models a protein using information from three different biological scales: the evolutionary scale, detailing the global evolutionary conservation of a residue in the species tree, its local conservation within a phylogenetic clade, and the epistatic contribution to its mutation; the molecular scale, focusing on the position of a residue within a structure and its physico-chemical properties; and the population scale, considering the allele frequency of a mutation in specific populations. Collectively, these factors enable PRESCOTT to optimally predict mutation effects and interpret variants.

Both PRESCOTT and AlphaMissense underscore the importance of structural information in mutation analysis. Key mutations are often located in protein binding sites (27–29) or in the structural core (25). More subtle long range allosteric effects in a structure have also been identified (48). In this regard, it should be noted that the structural component of the AlphaMissense predictor does not currently take into account the fact that most proteins are assembled into complexes with diverse quaternary structures, thus missing important information about how monomers interact. Future improvements on the protein assembly problem, will pave the way for new improvements of AlphaMissense (49). ESCOTT and PRESCOTT, for their part, utilise existing structural models to evaluate whether a residue is buried or part of a coiled region, focusing on residues hidden in the structural core or potential binding sites. While integrating precise protein binding site predictions could enhance ESCOTT’s mutational effect assessments, we have chosen to maintain model simplicity for biologically justifiable improvements. Additionally, ESCOTT and PRESCOTT process unstructured protein regions using evolutionary information, evaluating residues in potentially disordered regions based on their evolutionary conservation (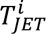). More sophisticated modelling of important residues present in intrinsically disordered regions could be incorporated into ESCOTT model to enhance predictions in these regions. These additions will be interpretable and useful for understanding the mechanical effects of mutations in proteins.

Population allele frequencies, a cornerstone in clinical diagnostics, are effectively utilized by both PRESCOTT and AlphaMissense. AlphaMissense is trained on allele frequencies and their known effects, whereas PRESCOTT integrates only allele frequency information to screen common variants into the final prediction stage, without relying on variant classification. PRESCOTT’s model, simple yet effective, combines ESCOTT scores based on evolution and structure with population frequencies, favoring specific human populations over a global approach (32). In doing so, it indirectly takes into account the genetic background that extends beyond the sequence of a protein and that depends on external factors, such as the environment and diet of an individual carrying the mutation. According to our calculations, this population-specific model provides slightly better estimates of the pathogenic versus the benign status of a mutation than a global population model. Given the importance of population allele counting in the analysis, the small number of different populations that are registered today, and the small number of individuals that have been genetically screened, broader exome sequencing efforts are essential for a deeper understanding of genetic diseases and, more generally, of protein behavior. Importantly, by providing both ESCOTT and PRESCOTT scores, we offer to geneticists the choice to either consider the ESCOTT score and use gnomAD or their allele frequency database of choice to make an expert classification or to directly rely on PRESCOTT for a fast and integrated prediction of variant pathogenicity.

Though ESCOTT and similar recent methods mark significant advancements in mutation effect prediction, their variable performance across different proteins – with Spearman correlation coefficients ranging from 0.135 for SCN5A to 0.690 for P53 – indicates that large-scale application is not yet feasible. Nevertheless, tools like PRESCOTT, AlphaMissense, EVE, ESM1b and all the others mentioned in this article will be invaluable for future variant prioritization. It should be noted, however, that PRESCOTT is uniquely designed for interpretability at each step, integrating data from evolutionary, molecular, and population scales. This multifaceted approach provides quantitative insights crucial for developing criteria to aid clinicians in making informed decisions. The cumulative effect of these components in PRESCOTT will offer substantial evidence to support clinical judgment, bridging the gap between genetic analysis and practical healthcare applications.

The integration of advanced models for handling allele frequency data is expected to enhance PRESCOTT. Both the MPC (Missense badness, PolyPhen-2, and Constraint) (50) and CCR (Constrained Coding Regions) (51) use population allele frequency to estimate average effect of missense variants in sub-genic regions. These models offer two distinct pathogenicity measures that help identify coding regions with a low frequency of missense variations. Such regions are known to contain a significant number of disease-related variants. These models are very interesting and differ significantly from PRESCOTT, which uses allele frequency information to exclude common variants, rather than for pathogenicity prediction like MPC and CCR. Implementing a strategy in PRESCOTT that focuses on regions with fewer pathogenic mutations, based on ESCOTT scores, could significantly enhance its capabilities. This strategy will be explored in future developments. Additionally, incorporating allele frequency data from both humans and primates (52) could further improve the results.

## ONLINE METHODS

### ProteinGym Dataset

We used the ProteinGym DMS dataset (www.proteingym.org/substitutions-dms-level) to investigate the agreement between ESCOTT predictions and experimental data. The dataset contains 87 DMS experiments performed on 72 proteins. Of these 87 experiments, 76 involve single point mutations only, for a total of 266,473 predictions, and the other 11 experiments contain 1,271,579 multiple point mutations. DMS was developed to systematically quantify the effects of genetic variations on a large scale, with high efficiency and relatively low cost (53,54). Fitness competition, toxicity, protein stability, binding affinity have been used to measure mutational effects for a protein-coding gene. The phenotypic values measured are then organized into large matrices of protein size per 20, i.e. the number of amino acids, reporting the measurements from each point mutation experiment.

### Multiple sequence alignments of ProteinGym Dataset

We obtained MSAs from Colabfold method using uniref30_2103 and colabfold_envdb_202108 sequence databases. This method produced MSAs for all proteins in the ProteinGym dataset without any problem and we use it as reference dataset for our analyses.

### Structures generated for ProteinGym and the Human Proteins Dataset

For proteins in the ProteinGym dataset, we computed model structures with Alphafold version 2.1.2 (41), except for three of the proteins, BRCA1 HUMAN, PABP YEAST, and SRC HUMAN, whose model structures have been obtained from the AlphaFold Protein Structure Database (55). Only the model with the highest rank was used as the reference structure. Since Alphafold implementation uses several databases such as Uniref90 (56), BFD (57,58) and Mgnify (59), due to their large sizes, Alphafold computational time becomes prohibitively high for thousands of proteins and, for this reason, we used Colabfold, based on highly clustered databases, to generate a single model structure for proteins in the Human Proteins Dataset.

### The Human Proteins Dataset is used to define a validation set and a training set

We used 3,013 clinically relevant human proteins to assess ESCOTT predictions against ClinVar labels. This dataset was used in (13) to evaluate performance of EVE method (60) against ClinVar labels (see below) and it was downloaded from evemodel.org<u>/download/bulk</u>. Structural models and MSAs have been generated for the entire set of 3,013 proteins using Colabfold (21,61–64). We calculated the entire single point mutational landscape for 3013 proteins (totalizing 33,771,588 missense variants) with ESCOTT using Colabfold MSAs, and conducted performance evaluation only on ClinVar mutations. Because of the comparison with EVE, ESM1b and AlphaMissense, we reduced the original number of 3,013 proteins considered by EVE to 2,383. The reduction is due to several reasons: 1. no benign and pathogenic labels were both available in ClinVar, 2. no direct matching between ensemble identifiers and gene names was available, 3. no analysis was present in the AlphaMissense dataset. Note that ESM1b was run by us on all 2,383 proteins.

For the purpose of calibrating default parameter values in ESCOTT, we randomly selected a subset of 500 proteins from our broader dataset of 2,383 proteins, ensuring that none of these proteins are members of the ACMG list. Specifically, this subset was instrumental in setting the parameters such as ‘frequencyCutoff’ and ‘scalingCoefficient’ for the PRESCOTTscore-low-scores algorithm (detailed further below), as well as establishing the two critical thresholds that distinguish benign, pathogenic, and VUS in the ESCOTT analysis. These thresholds were precisely calculated using data from 1,849 pathogenic mutations and 2,523 benign mutations, each linked to the aforementioned 500 proteins as classified by ClinVar. To ensure the integrity of our validation process, we excluded these 500 proteins from our final test dataset. Consequently, our validations were rigorously conducted on a separate group of 1,883 human proteins. The specifics of the proteins in these two distinct datasets are available in our supplementary data files for reference.

### ACMG Actionable Genes Dataset (v3.1)

We considered the full set of 78 actionable genes from version 3.1 of the American College of Medical Genetics and Genomics (ACMG) dataset (42). The TTN_HUMAN protein was too big to be modeled and was excluded; the ACVL1_HUMAN protein was excluded because it did not belong to the EVE dataset; 12 more proteins were discharged because they had neither benign nor pathogenic labels. We remained with 64 proteins. We modeled structures of 61 of them with Colabfold (43–45,64) and Alphafold (47). For the remaining three (RYR1_HUMAN, RYR2_HUMAN and APOB_HUMAN) no full structural model existed.

#### A new dataset of gain-of-function mutations

We focused our study on 13 genes known implicated in autoinflammatory diseases: *ALPK1, CDC42, MEFV, PSTPIP1, NLRC4, NLRP1, NOD2, CEBPE, RIPK1, PLCG2, CARD14, STING*, and *IFIH1*. These genes were carefully selected based on information from Infevers (65), augmented by thorough reviews of relevant literature and insights from unpublished experimental data. Our objective was to meticulously identify and select a dataset comprising over 100 missense pathogenic gain-of-function mutations, ensuring high confidence in their pathogenicity.

#### The Comprehensive Human Protein Database in ESCOTT webserver

The ESCOTT webserver features precomputed ESCOTT analyses for 19,432 human proteins, with model structures sourced from the AlphaFold Human Proteome Structures Database (AFDB). In addition to this, the server provides iGEMME analysis results for these proteins. However, it is important to note that AFDB does not include model structures for proteins exceeding 2700 amino acids in length, and therefore, their analyses are not available on the ESCOTT server. For analyzing these larger proteins, users are advised to employ Colabfold to create MSAs and generate structures of lower accuracy for ESCOTT server analysis. It should also be noted that eight human proteins—APC, FBN1, BRCA2, FLNC, DESP, RYR1, RYR2, APOB—listed in the ACMG dataset are excluded from the ESCOTT database due to their length. Their analyses are instead provided in the dedicated ACMG folder.

Regarding the 19,432 proteins with ESCOTT output files, there was one failure involving a very short protein (UniProt ID A0A075B6Y3). In addition, 14 proteins (UniProt IDs: 5SY13, Q75NE6, Q9BZK8, Q0P140, Q9H2U6, Q9Y6C7, Q13278, Q9HBX3, Q8TCH9, Q6ZQT0, Q9UJ94, Q9BYR3, Q9BQ66, A8MUU9) have incomplete ESCOTT output files, missing scores in some MSA regions. Consequently, complete analyses are available for 19,417 proteins.

### The gnomAD database

The Genome Aggregation Database (gnomAD) is currently the largest and most widely used publicly available collection of population variation from harmonized sequencing data (https://gnomad.broadinstitute.org/news/2023-11-gnomad-v4-0/) enabling variant analysis (32). The gnomAD dataset is used in the majority of rare disease analysis pipelines in both diagnostic and research settings around the world. gnomAD follows the rigorous guidance for the evaluation and aggregation of variant evidence determined by ACMG and Association for Molecular Pathology (AMP) (66). There are three available versions, gnomAD v2.1, which contains data from 125,748 exomes and 15,708 whole genomes, all mapped to the GRCh37/hg19 reference sequence, gnomAD v3.1, which contains 76,156 whole genomes (and no exomes), all mapped to the GRCh38 reference sequence, and gnomAD v4.0.0, which is nearly five times larger than the combined v2/v3 releases and consists of exome sequencing data from 730,947 individuals, and genome sequencing data from 76,215 individuals. Both callsets were aligned to build GRCh38 of the human reference genome. The results reported in this study refer to gnomAD v4, which includes a higher number of coding variants compared to the previous versions. It covers data from 8 populations: gnomAD collects data from eight different populations, African/African American, Admixed American, Ashkenazi Jewish, East Asian, European (Finnish), European (non-Finnish), South Asian, and Middle Eastern. gnomAD uses the ClinVar classification of variants into five categories: benign, likely benign, uncertain significance, likely pathogenic, and pathogenic. PRESCOTT has been run on the 1,883 distinct proteins in gnomAD, comprising 7,954 pathogenic mutations and 9,276 benign ones (**Table 2**). These mutations are associated to their population origin. To compare PRESCOTT performance to EVE performance, we considered ClinVar labels from gnomAD and searched for the corresponding protein in the EVE data. If EVE computed mutational values for that mutation, we considered it.

**Table 2.** gnomAD v 4.0 (November 2023) database characteristics.

| Stars | Distinct proteins | Pathogenic | Benign |
| --- | --- | --- | --- |
| ≥0 | 1,883 | 7,954 | 9,276 |
| ≥1 | 1,761 | 5,351 | 9,112 |
| ≥2 | 1,133 | 2,509 | 4,039 |
| ≥3 | 22 | 335 | 365 |

### The ClinVar dataset and its stars subsets

The ClinVar database includes germline and somatic variants of any size, type or genomic location. Interpretations are submitted by clinical testing laboratories, research laboratories, locus-specific databases, expert panels and practice guidelines (67,68). gnomAD data are annotated by ClinVar labels as VUS, pathogenic or benign. Precisely, ClinVar labels ‘Pathogenic’, ‘Pathogenic/Likely pathogenic’ and ‘Likely pathogenic’ were employed to define the set of pathogenic mutations in PRESCOTT. Similarly, we selected ‘Benign’, ‘Benign/Likely benign’ and ‘Likely benign’ labels to form the dataset of benign mutations. Also, the mutations reported in ClinVar contain information about review status of each mutation (www.ncbi.nlm.nih.gov/clinvar/docs/review_status/), expressed with an integer number of ‘gold stars’ indicating the number of sources that verified the effect of the mutation: 0 stars indicate mutations submitted without interpretation; 1 star indicates mutations provided by one submitter with interpretation and evidence; 2 stars indicate mutations submitted by two or more labs with the same interpretation; 3 stars indicate a review from external panels. The full ClinVar dataset has been referred to as “star 0”, indicating the fact that many alleles have been reported without interpretation. Three smaller datasets, “star 1”, “star 2” and “star 3” are defined inclusively, e.g. “star 2” includes all mutations labelled with 2 and 3 stars. The mutations in gnomAD labelled with ClinVar annotations used in our analysis are described in **Table 2**.

Note that the amino acid mutations reported in ClinVar stand one nucleotide position away from the amino acid reported in the reference human protein. This means that instead of the full ESCOTT predicted matrix, PRESCOTT evaluates only some of the positions of the matrix, namely those that are supposed to appear in human populations, possibly causing diseases. The November 2021 ClinVar report on these mutations is used in our analysis as ground truth, as we need to compare our results with EVE’s calculations.

### Details on the ESCOTT model

ESCOTT relies on two main important modelling steps. The first defines how to represent the residues at a specific position in the query sequence and the second estimates the mutational effect of a variant at that position.

#### Modelling residues in the query sequence

To each position i in a query q, ESCOTT provides a score to the residue at that position by considering information coming from both evolution and structure, whenever possible. For this, positions are first categorised in two classes, as either participating to the structure or remaining unstructured. In the first case, the position is modelled by the highest value among two terms, one modelling residues which lie potentially at the interface of the protein with other molecules, and another one modelling residues in the protein structural core:

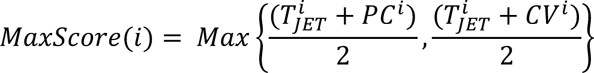

where i indicates the amino acid position in the protein. The first term in the *MaxScore* formula models residues that are expected to be evolutionary conserved (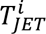, Figure 1C) (23) and to satisfy physico-chemical properties at a binding site (PC^i^). The second term models residues that are expected to be evolutionary conserved (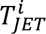) and to lie at the structural core of the protein (CV^i^). Note that there is no explicit prediction of residues lying at the interfaces but merely a modelling of residues with conserved physico-chemical properties, expected at the interfaces, indicating a potential interaction with other molecules (24). The maximum between the value of the two terms selects the effective structural role of the position. When a residue in the sequence belongs to a coiled region, ESCOTT uses only its evolutionary conservation 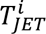 to model it, based on the intuition that if a residue is evolutionary conserved, it should be important and hence susceptible to mutations. Note that T_JET_, PC and CV values fall in the [0-1] range.

#### Modelling the mutational effects of a variant

This model is based on GEMME (20) architecture, where the usage of T_JET_ is replaced by the new modelling of the residues in a query sequence described above. We remind, for clarity, the basic terms of the model which is fully described in (20). For each position i and each mutation X-to-Y in a query sequence q, ESCOTT defines a predicted effect (PE) of the mutation as a combination of an independent (Ind) and an epistatic (Epi) term linearly combined with a scalar coefficient

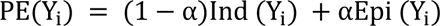

where α is a linear coefficient set to 0.6 by default, giving more weight to the epistatic contribution of the model. Intuitively, Ind (Y_i_) describes the amino acid frequency observed at a given position independently from other positions in the sequence, and it is computed as the relative frequencies of Y_i_ (within a reduced alphabet (20)). Epi(Y_i_) uses the *MaxScore* at position i, and models the epistatic contribution by taking into account all positions in the sequence that need to be changed for the mutation to be accepted (Figure 1F; equation 4 in (20)). The PE(Y_%_) scores are finally normalized (Figure 1E) with *MaxScore* to obtain a normalized predicted effect (NPE; equation 8 in (20); Figure 1G):

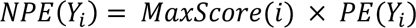

### PRESCOTT model of population frequency in gnomAD

For the analysis of human mutations, PRESCOTT considers the 8 populations in gnomAD independently and establishes, for each population, the PRESCOTT score by integrating to the ESCOTT score a term which is dependent on a popmax gnomAD population frequency. This is done with the following algorithm depending on two parameters, the “frequencyCutoff” and the “scalingCoefficient”:

#### Algorithm PRESCOTTscore-low-scores

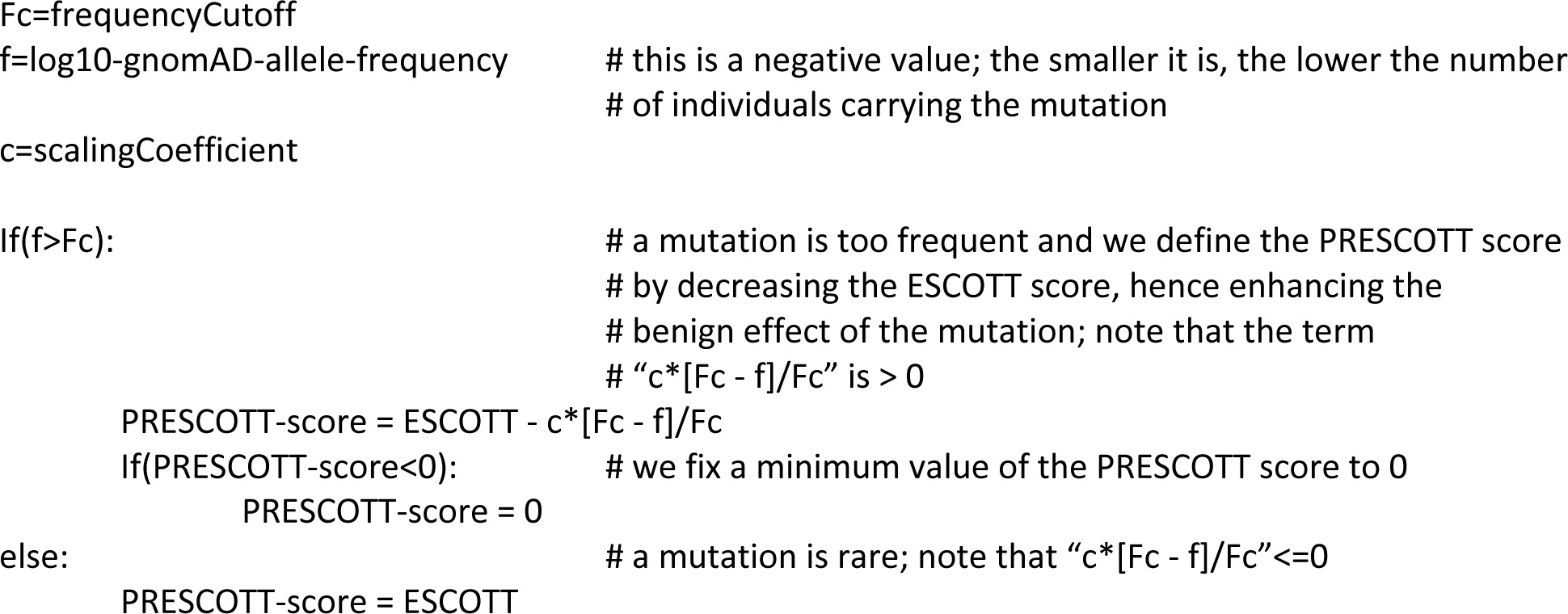

with default parameter values frequencyCutoff=-4 (where −4= log_10_(0.0001)) and scalingCoefficient=1. They have been determined based on best performance over the eight populations of the gnomAD v4.0.0 dataset (see **Figure S8A**) and the training set of 500 proteins defined from the Human Protein dataset.

All tests reported in the article have been realized with these default values. After evaluating a mutation on each population, PRESCOTT chooses the popmax allele frequency, defined as the population maximum allele frequency in the continental populations (African/African American, Admixed American, Ashkenazi Jewish, East Asian, European/Finnish, European/non-Finnish, South Asian, and Middle Eastern) as recommended in (32). Generally, if a variant is common in one population, it can be assumed to be benign across all populations. Based on this idea, the algorithm enhances a PRESCOTT value towards 0 by calibrating it towards a “benign” prediction.

A symmetric treatment of high scores could enhance the prediction of pathogenic mutations. For this, we defined the following algorithm:

#### Algorithm PRESCOTTscore-low-and-high-scores

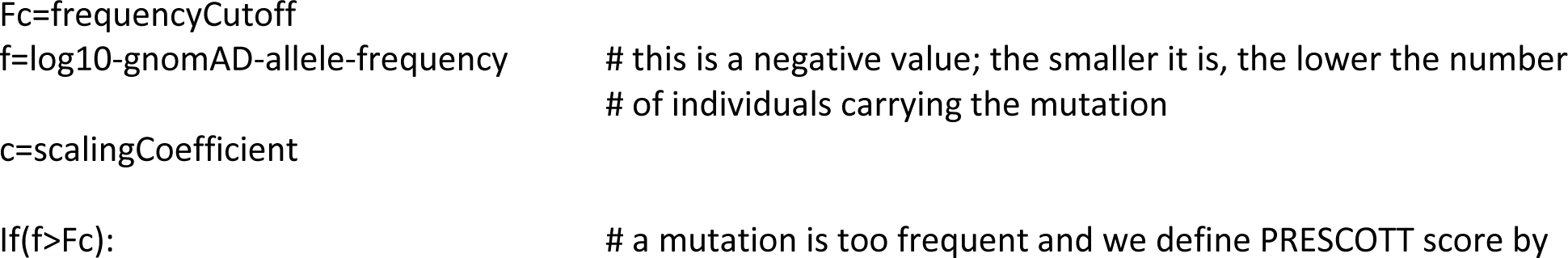
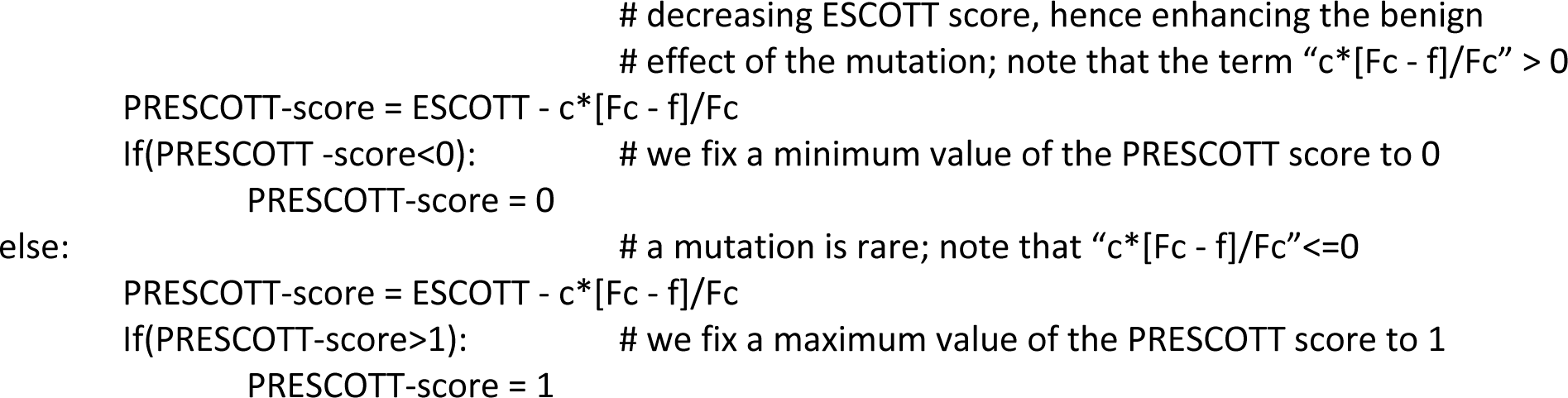

with default parameter values frequencyCutoff=-3.5 and scalingCoefficient=0.75, determined on a best performance on the gnomAD database (**Figure S8B**) and the training set of 500 proteins defined from the Human Protein dataset. The idea of this algorithm is that a mutation that is frequent in some population is most probably “benign” and, for this, it reduces the score, as in **PRESCOTTscore-low-scores**. In contrast, if a mutation is rare in all populations, it enhances ESCOTT value towards a “pathogenic” prediction by augmenting the PRESCOTT value towards 1. Note that the lack of information in gnomAD due to a relatively small number of populations and individuals sequenced, introduces biases towards pathogenic diagnoses which will improve in the future, with higher population sampling. In **Figure S8B**, one can clearly see a small number of benign mutations that end-up with high PRESCOTT scores. This effect could be undesirable for doctors, but interesting for suggesting candidates for experimentation. We believe that sufficient data are not yet present in public databases to safely produce predictions based on this hypothesis. The algorithm is implemented in the PRESCOTT package but it not used as a default method.

The frequency of gnomAD alleles in the Star >0 set, considering all populations at once, was tested, but performance was slightly inferior (0.94).

### The ESCOTT model incorporates secondary structure information and knowledge of the protein core

We used Biotite Python package (69) and DSSP (70) to assign secondary structure of each amino acid in the protein structures. In ESCOTT, after the secondary structure assignments, we selected *MaxScore* when an amino acid is in any one of these secondary structural elements: H, E, G, B, I, S and T. When the amino acid is in a coiled region, we performed a conditional selection. We selected *MaxScore* if the coil is shorter than 5 amino acids and we selected T_&’(_ score for all the other coils. This approach helped us to distinguish coils linking secondary structural elements such as helices or strands, from coils in disordered regions. Even if slightly different coil lengths are selected (4–6), our method produces similar results (not shown).

To compute whether a residue belongs or not to the protein core, we use its circular variance (CV), a measure of the vectorial distribution of a set of neighbouring points around a fixed point in 3D space (26,71). For a given residue, CV reflects the density of the protein around it. The CV value of an atom *i* is computed as:

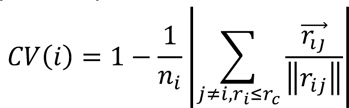

where *n_i_* is the number of atoms distant by less than r_c_ Å from atom *i*. The CV value of a residue *j* is then computed as the average of the atomic CVs, over all the atoms of *j*. A low CV value indicates for a residue that it is located in a protruding region of the protein surface. CV values are scaled between 0 (most protruding residue of the protein) and 1 (least protruding residue of the protein) for the calculation of residue scores. The value r_c_ is set to 7Å after an evaluation on best performance over the set of DMS experiments (**Figure S9**). This default value has been used in all validation testing reported and on the predictions of the 19000 proteins.

An evaluation of the predictions on structured versus coiled regions along the sequence is reported in **Figure S2** for all proteins in the ProteinGym dataset. As expected, the analysis shows that predictions are more precise on structured regions.

### The ESCOTT model incorporates physico-chemical propensities of amino acids at the interface

Physico-chemical properties (PC) of amino acids are derived from propensities specific to every amino acid to be located at a protein interface, taken from (30). The original values, ranging from 0 to 2.21, are scaled between 0 and 1 for the calculation of residue scores.

### Analysis of the independent and epistatic modules building ESCOTT

ESCOTT is defined on the contribution of its independent (Ind (Y_%_)) and epistatic (Epi (Y_%_)) terms, evaluating the mutation from X_i_-to-Y_i_. To evaluate the importance of these contributions, we considered ESCOTT performance on the reference ProteinGym dataset, constituted by 87 DMS experiments performed over 72 different proteins. ESCOTT average Spearman correlation on this dataset is 0.492. The average Spearman correlation of the independent model, taken alone, is 0.459 and of the epistatic one, taken alone, is 0.465. A detailed description of the correlations for each protein is reported in **Figure S10** showing that the combination of the two terms is indeed necessary to sensibly improve the performance for almost all proteins. Moreover, on most proteins, the epistatic model alone performs better than the independent one as already observed in (20).

### ESCOTT performance and impact of the alignments in ProteinGym

Changes in the underlying sequence datasets representing the input proteins can induce slight improvements in performance. We considered Uniref100 and (the highly clustered) Colabfold (21,61–64) databases and obtained average Spearman correlations of 0.489 and 0.492 for the two datasets, respectively. To guarantee a fair comparison of PRESCOTT with other approaches, we consider datasets of sequences based on Colabfold.

### The ESCOTT program is built on iGEMME

ESCOTT has been developed from the improved version iGEMME of GEMME. In iGEMME, the characteristics of the input sequence dataset and default parameters are optimized. When a user does not provide an MSA, the default method to obtain MSAs in the GEMME webserver is PSI-BLAST (*72,73*). In ESCOTT, we use the Colabfold method (*64*) due to its efficiency and clustering procedure. In addition, the maximum number of sequences processed has been increased from 20000 to 40000. A parameter that restricts the number of sequences due to computational load (CPU load or max_load parameter) was increased from 500,000 to 8,000,000. Also, the maximum heap size in the calls of the JET program was increased from 1,024MB to 8,192MB. These parameters’ changes helped us to handle large proteins and MSA files containing thousands of sequences in ESCOTT. This improved version of GEMME, called iGEMME, based on the new parameterization, is the nutshell of the ESCOTT program. **Figure S11** provides a comparison of ESCOTT with iGEMME and GEMME. The iGEMME average Spearman over the entire ProteinGym dataset of 0.472 (red, **Figure S4**) already improves GEMME performance (0.464; violet, **Figure S11**).

#### Comparison with EVE, ESM1b, and AlphaMissense

EVE results on the 87 ProteinGym experiments and on the 3013 human proteins were downloaded from www.proteingym.org/substitutions-dms-level and evemodel.org<u>/download/bulk</u>, respectively. Note that the label “EVE (ensemble)” for the ProteinGym dataset indicate that EVE scores were calculated from the approximate EVE posterior distribution by ensembling scores over 5 independently trained variational autoencoders (see Extended Data Fig. 2 in (60)). EVE (ensemble) was used for all 87, single and multiple, mutation experiments. For the ∼3000 proteins, EVE(ASM) results were used. On the ProteinGym experiments, performance of all other tools cited here are taken from www.proteingym.org/substitutions-dms-level. ESM1b was downloaded from <u>github.com/ntranoslab/esm-variants</u> and run on all single point mutation predictions for 3013 proteins (on a Dell Precision 7920 Workstation with 40 CPUs and 64 GB RAM).

#### Computational time performance of ESCOTT on the ProteinGym dataset

The implementation of the ESCOTT model is computationally very efficient. All proteins in the ProteinGym dataset of about 1.5 million mutations can be completed within 6.16 hours (**Figure S12**). The most time consuming experiments are the highly dense HIS7_YEAST_Pokusaeva_2019, which contains 496137 multiple point mutations (74) processed with a total computing time of almost an hour. HIS7_YEAST is 220 amino acid long and the MSA file used for the predictions contains a large number of sequences (26,706). Predictions for SCN5A_HUMAN_Glazer_2019 also took about one hour due to large size of the protein (2016 amino acids) and numerous homologs (19760). Calculations for all other experiments could be completed within a few minutes and average computing time of 255.1 seconds per experiment. Calculations were performed on a workstation with 40 core Intel(R) Xeon(R) Silver 4210R @ 2.40GHz CPUs and 64 GB RAM. Note that graphical processing units are not needed for PRESCOTT calculations. EVE computational time has been already evaluated in (20): EVE computing time is a few orders of magnitude higher than iGEMME and ESCOTT methods.

#### ESCOTT ranksorted scores

A direct comparison with EVE on the Human Protein dataset and ClinVar dataset asks for a normalization of ESCOTT scores in the range [0,1]. ESCOTT scores in entire ESCOTT matrices has been normalized with ranksorting, using the command line “from scipy.stats import rankdata”. After the data is ranked, score positions in the ranked list are divided by the number of datapoints in the list to make values ranging in [0,1]. Due to the direct comparison with EVE scoring, we reversed the ranksorted scores so that values close to 1 will be intended as “pathogenic” and those close to 0 as “benign”. Note that ESCOTT (and iGEMME) low raw scores, approximately varying from −10 to −4, correspond to pathogenic mutations. The use of ranksorting is indicated in all corresponding analysis.

It’s important to mention that rank sorting is applied to individual genes or proteins. In fact, the ESCOTT model employs evolutionary measures that differ across proteins, identifying some as less conserved and others as highly conserved. This leads to an internal re-evaluation of the score values assigned to each residue within a protein. Consequently, such reassessment amplifies the variations in score values within the protein itself.

#### Software

PRESCOTT source code is available. iGEMME and ESCOTT are a part of the PRESCOTT software. The user should provide an alignment file (mandatory argument and a pdb file (optional argument) as input. If a pdb file is not provided, ESCOTT model evaluates the role of each residue in a sequence by considering its evolutionary conservation (T_JET_) as the default feature for the amino acid. Otherwise, it uses the two-component max value terms, respectively combining circular variance, CV, and physico-chemical properties, PC, with evolutionary conservation. As a result, complete matrices describing mutational effects are provided, even in absence of a structural model. In this case, they will correspond to iGEMME.

#### Webserver

PRESCOTT webserver is accessible at the link - COMING SOON.

## Data Availability

All data produced are available online at
All the data generated in this work is provided in dedicated Zenodo repositories. ESCOTT predictions for the 3013 human proteins of EVE dataset: https://doi.org/10.5281/zenodo.10435332. ESCOTT predictions for the ProteinGym Dataset: https://zenodo.org/doi/10.5281/zenodo.10419509. ESCOTT predictions for the Human Proteome dataset of more than 19,000 proteins: https://doi.org/10.5281/zenodo.10577421. iGEMME predictions of the Human Proteome dataset of more than 19,000 proteins: https://doi.org/10.5281/zenodo.10441521. The gain-of-function ESCOTT single point mutational data was extracted from the Human Proteome dataset in https://doi.org/10.5281/zenodo.10442849. The ACMG ESCOTT data was extracted from: https://doi.org/10.5281/zenodo.10438640.

## Acknowledgments

This work was granted access to the Jean Zay HPC resources of IDRIS (www.idris.fr/eng/index.html) under the allocation 2022-AD010713586 made by GENCI. We extend our sincere gratitude to Francesco Oteri for his invaluable assistance in setting up the webserver.

## Author contributions

AC conceptualized the study with input from MT and TH; AC managed and supervised the project; MT developed ESCOTT and PRESCOTT software; MT performed experiments with the help of AC; AC and MT analyzed data and prepared figures; LD implemented the PRESCOTT webserver; AC wrote the manuscript with input from MT. All authors reviewed the manuscript.

## Data and material availability

All the data generated in this work is provided in dedicated Zenodo repositories. ESCOTT predictions for the 3013 human proteins of EVE dataset: https://doi.org/10.5281/zenodo.10435332. ESCOTT predictions for the ProteinGym Dataset: https://zenodo.org/doi/10.5281/zenodo.10419509. ESCOTT predictions for the Human Proteome dataset of more than 19,000 proteins: https://doi.org/10.5281/zenodo.10577421. iGEMME predictions of the Human Proteome dataset of more than 19,000 proteins: https://doi.org/10.5281/zenodo.10441521. The gain-of-function ESCOTT single point mutational data was extracted from the Human Proteome dataset in https://doi.org/10.5281/zenodo.10442849. The ACMG ESCOTT data was extracted from: https://doi.org/10.5281/zenodo.10438640.

## Software availability

PRESCOTT source code is available at http://gitlab.lcqb.upmc.fr/tekpinar/PRESCOTT/. iGEMME and ESCOTT are a part of the PRESCOTT software. We provide a docker image of PRESCOTT at https://hub.docker.com/repository/docker/tekpinar/PRESCOTT-docker/general.

---

**Figure S1.**
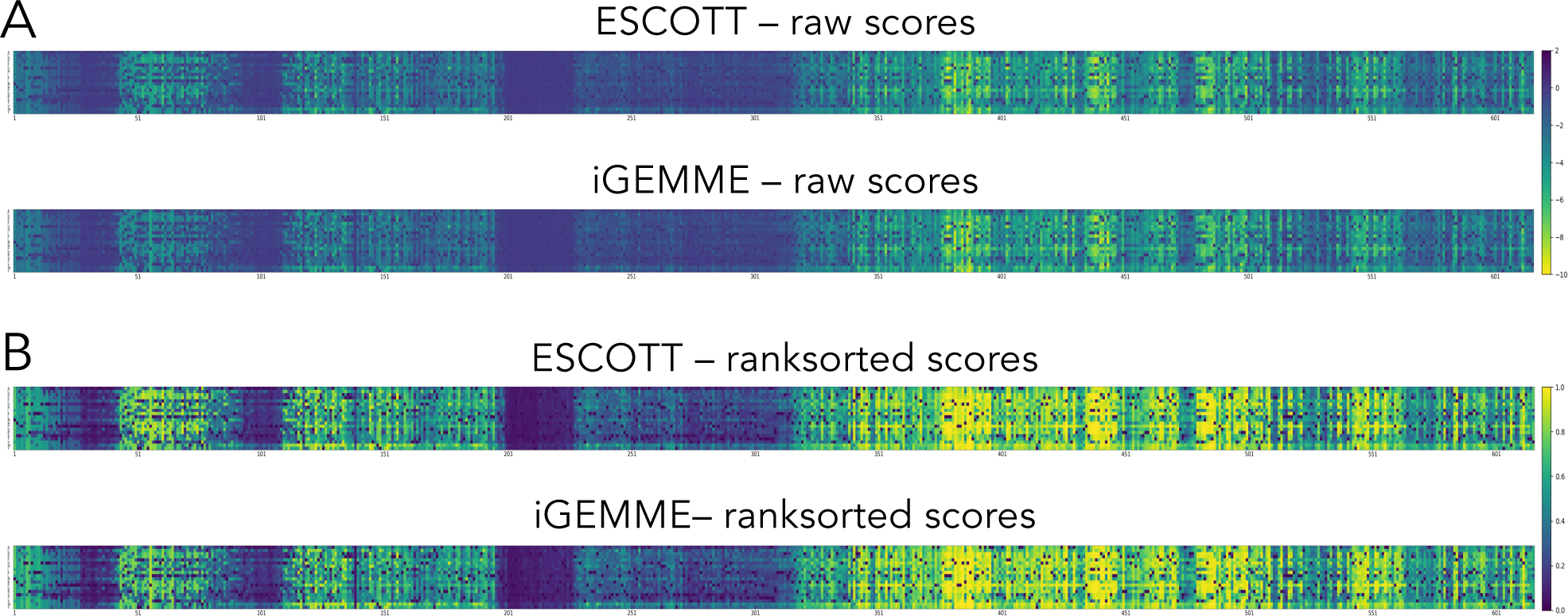
ESCOTT and iGEMME mutational matrices of the spastin protein. Comparison between raw (A) versus ranksorted (B) scores for the spastin protein full length predictions. Raw ESCOTT and iGEMME scores vary from −10 to 2 and ranksorted scores vary from 0 to 1. iGEMME and ESCOTT highlight essentially the same sensitive (lighter) regions of the protein. Ranksorted scores better emphasize these regions, with more contrasted score values for ESCOTT compared to iGEMME.

**Figure S2.**
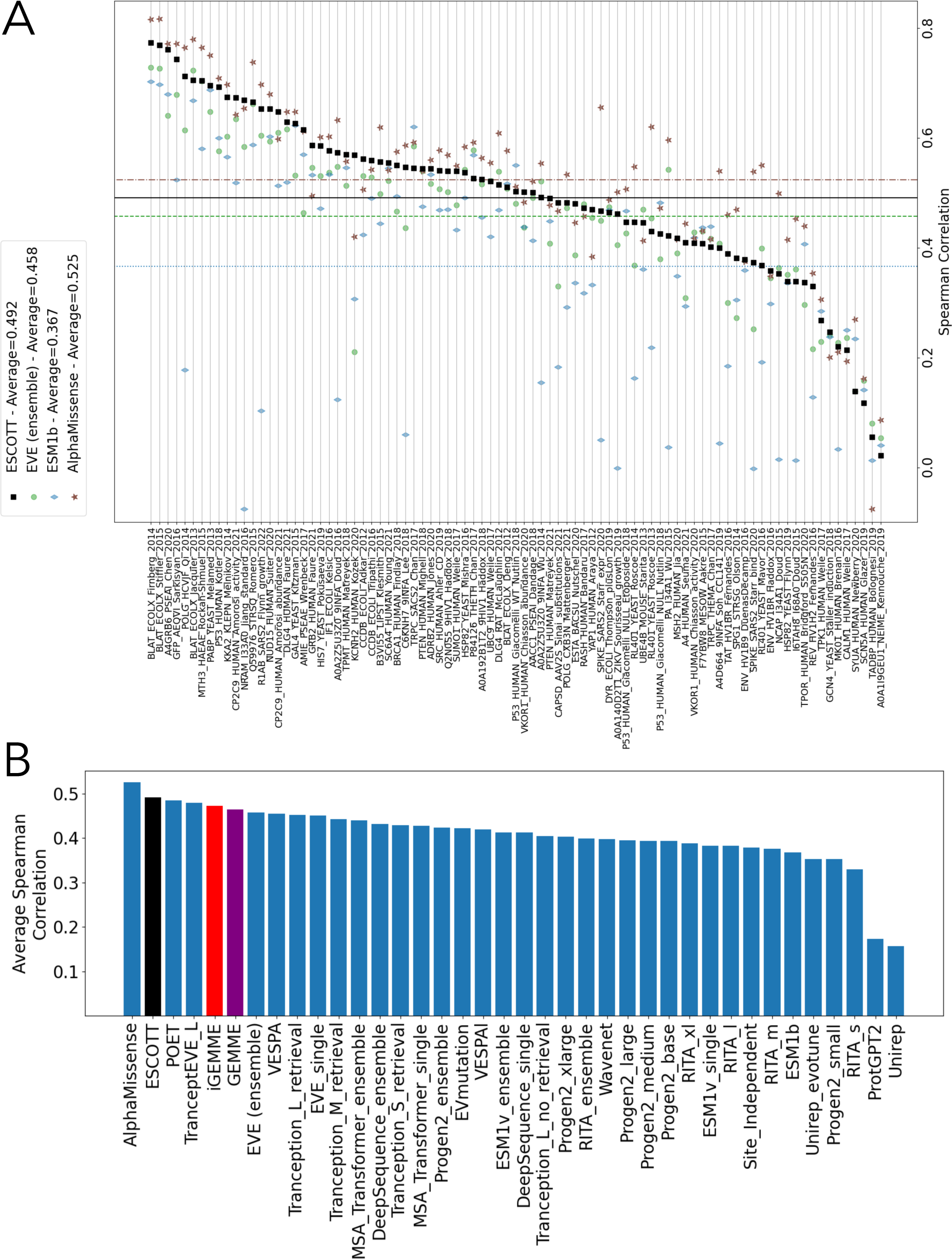
Comparison of ESCOTT, AlphaMissense, EVE (ensemble), ESM1b, and other methods on 87 deep mutational scanning experiments. **A.** ESCOTT (black stars), EVE (ensemble, green circle), ESMb1 (blue diamond), and AlphaMissense (brown square) Spearman correlation coefficients of the predictions with experimental data from the 87 Deep Mutational Scanning experiments of the ProteinGym dataset. Lines indicate averages: solid black line for ESCOTT, dashed dotted brown line for AlphaMissense, dashed green line for EVE (ensemble), dotted blue line for ESM1b. **B.** Comparison of ESCOTT with existing methods. The list of methods includes EVE (ensemble), AlphaMissense, GEMME, and ESM1b. ESCOTT is black, iGEMME is red and GEMME is purple. Data for all other methods was obtained from https://www.proteingym.org/substitutions-dms-level, except for AlphaMissense and PoET which are reported from (18) and (77) respectively. TranscriptEVE_L code was not available at the time of writing.

**Figure S3.**
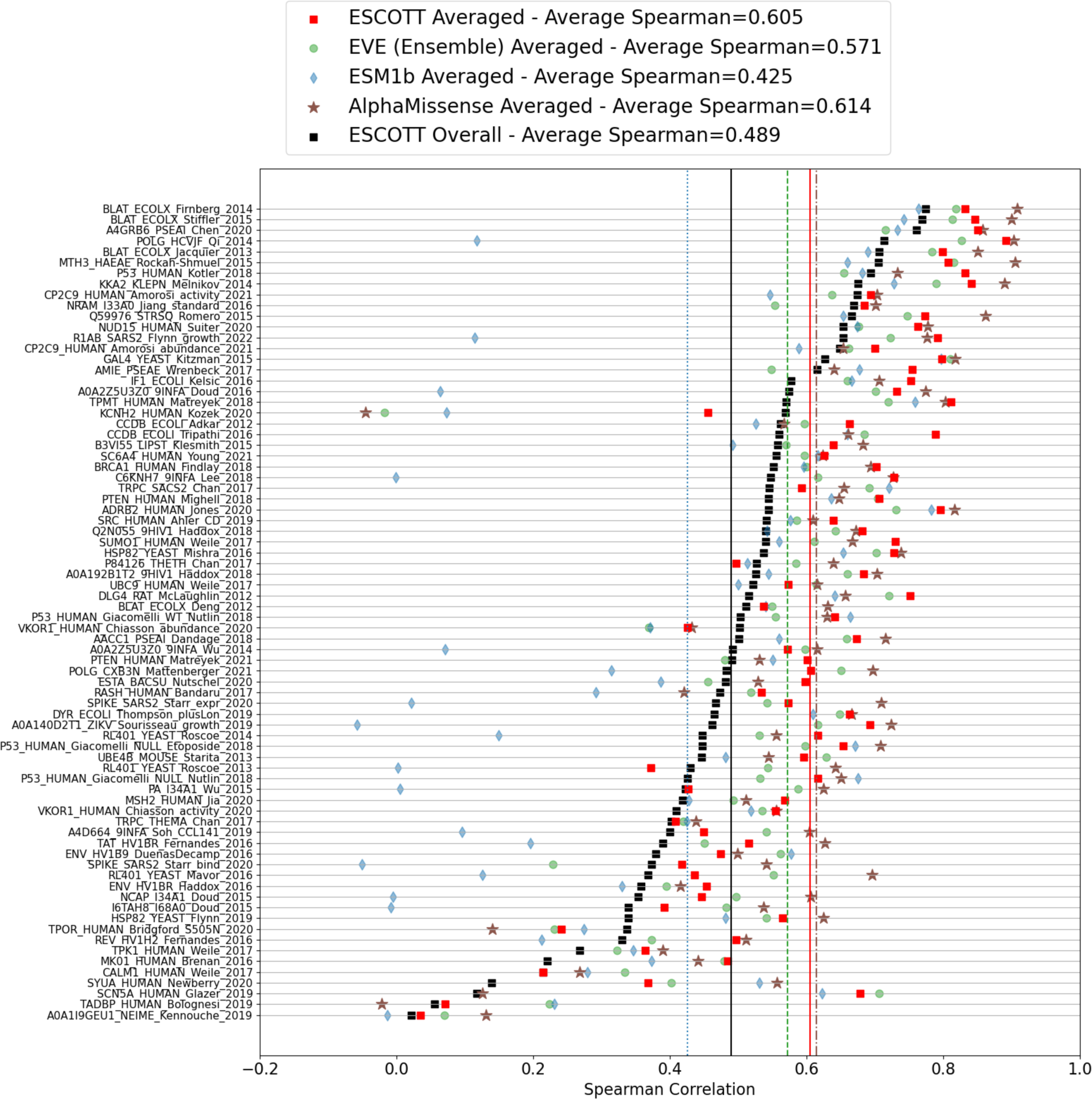
Spearman correlation coefficients for proteins in the ProteinGym dataset based on positional average scores: comparison of ESCOTT, EVE, ESM1b and AlphaMissense. Spearman correlation coefficients computed for the ESCOTT score matrices and the corresponding DMS matrices (black) versus positional averages (red). Only proteins of the ProteinGym dataset with single mutation experiments are considered. Black squares correspond to the values shown in **Figure S2** for ESCOTT. Performance of EVE (ensemble, green circle), ESMb1 (blue diamond), and AlphaMissense (brown stars) is also shown.

**Figure S4.**
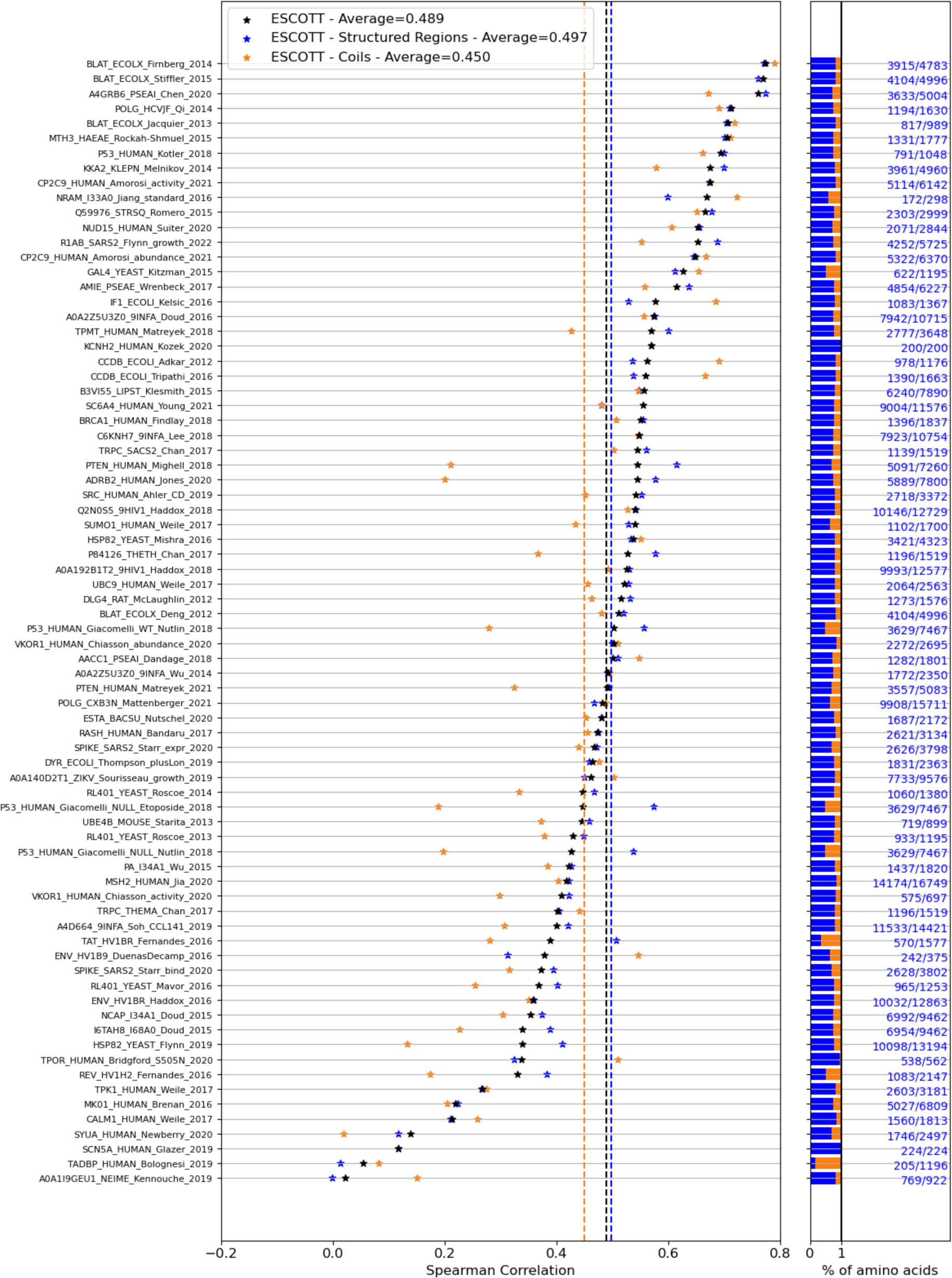
Comparison of ESCOTT for all mutations (black) versus mutations located in structured (blue) and coiled (orange) regions. Experimental measures and PRESCOTT scores are considered for the 76 **single point mutation experiments** in ProteinGym. Structured regions include H, E, G, B, I, S and T structural types as described in DSSP notation. Horizontal bars on the right show the percentage of mutations located in structured (blue) and coiled (orange) regions, for each protein. Note that some positions in a protein might have been discarded from counting because no experiment was reported on it. Dashed lines indicate averages.

**Figure S5.**
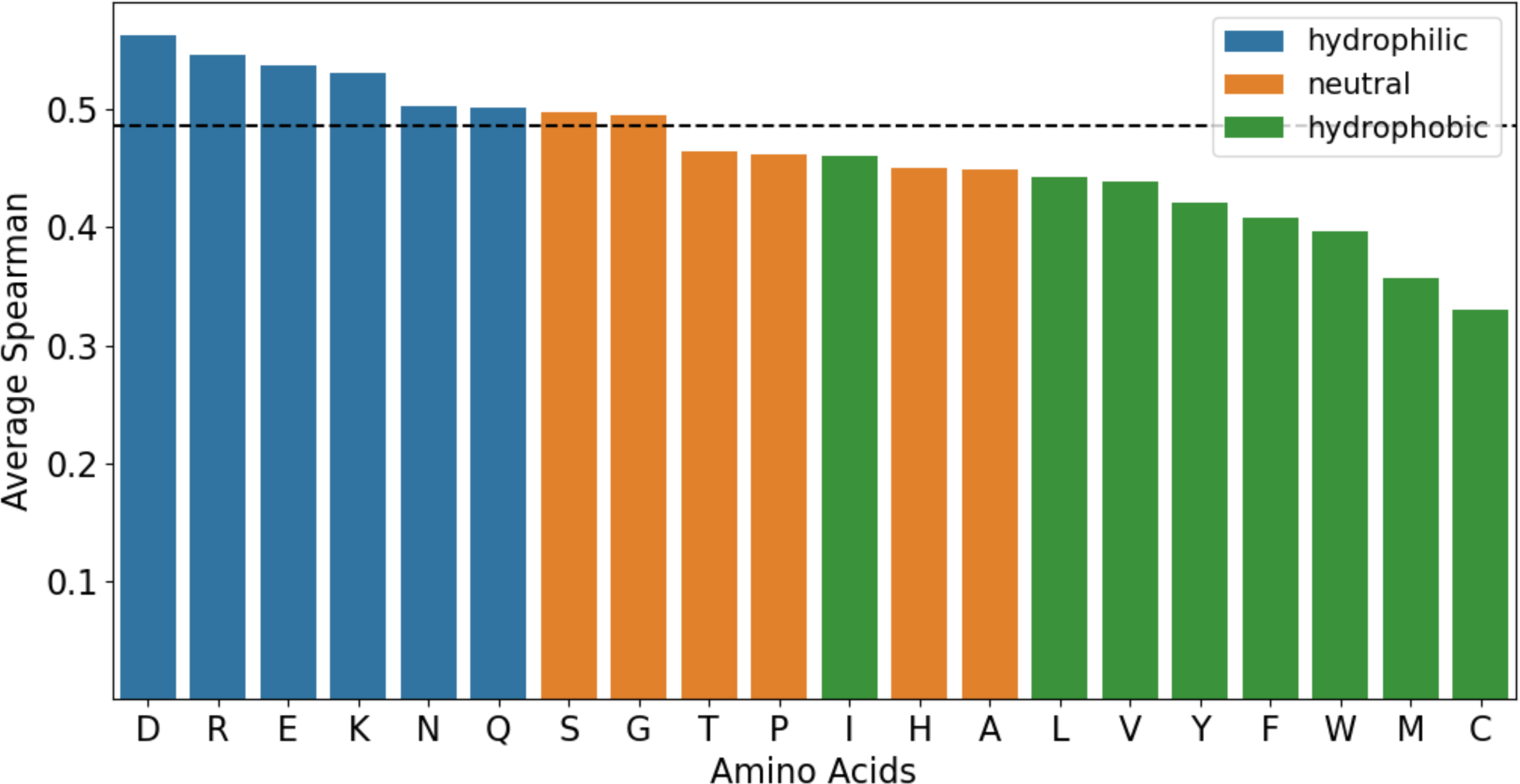
Average Spearman correlation for mutations towards the 20 specific amino acids. The amino acid bars are colored according to hydrophobicity. The dashed line indicates the average Spearman correlation coefficient for ESCOTT (0.485) computed over the 76 single point mutational experiments. Colors follow the amino acid three-class partition by hydrophobicity used in https://weblogo.threeplusone.com/manual.html.

**Figure S6.**
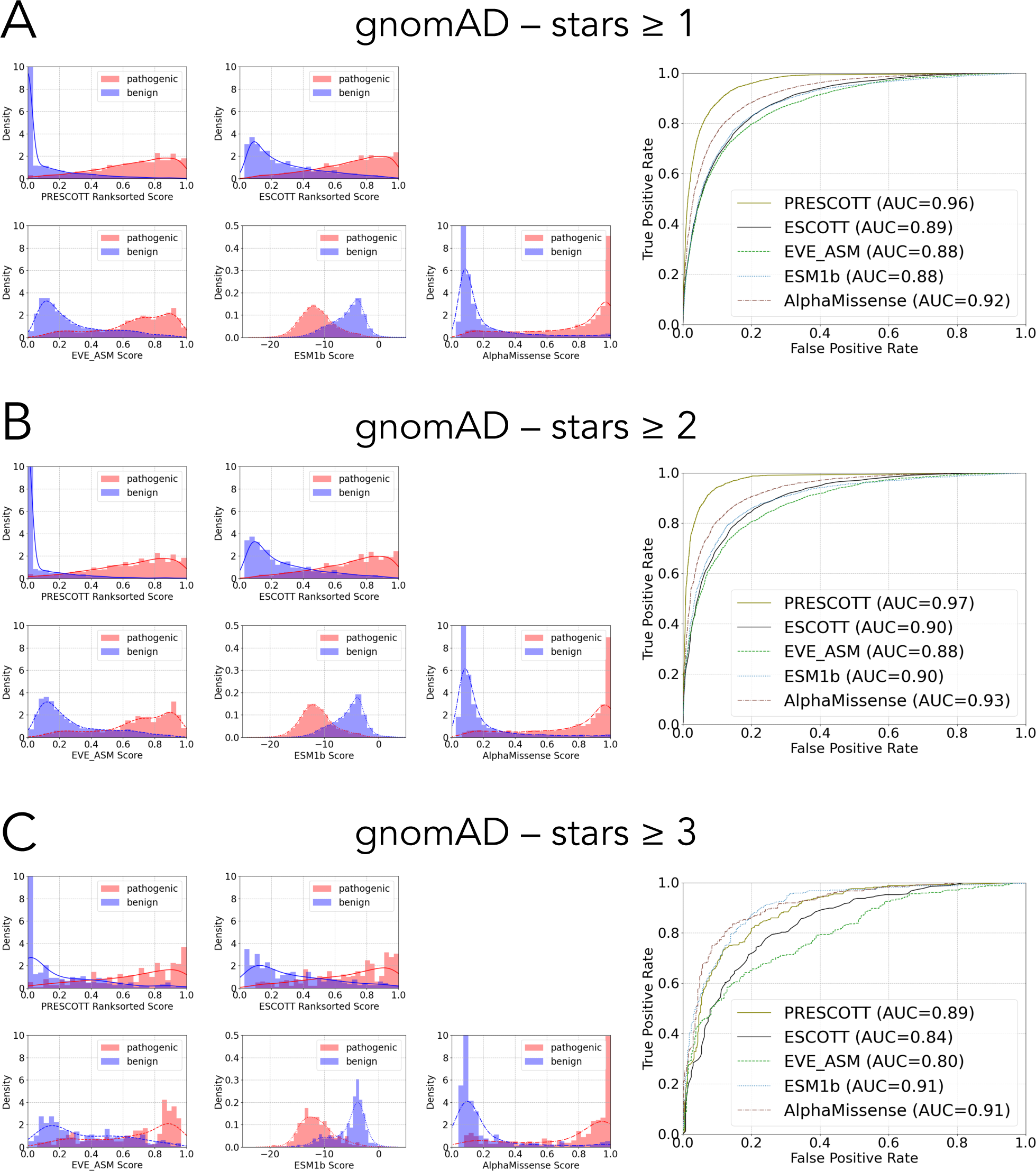
PRESCOTT performance on the dataset of human proteins informed by gnomAD v4.0.0 population frequencies for different review statuses (stars). Compare to Figure 7.

**Figure S7.**
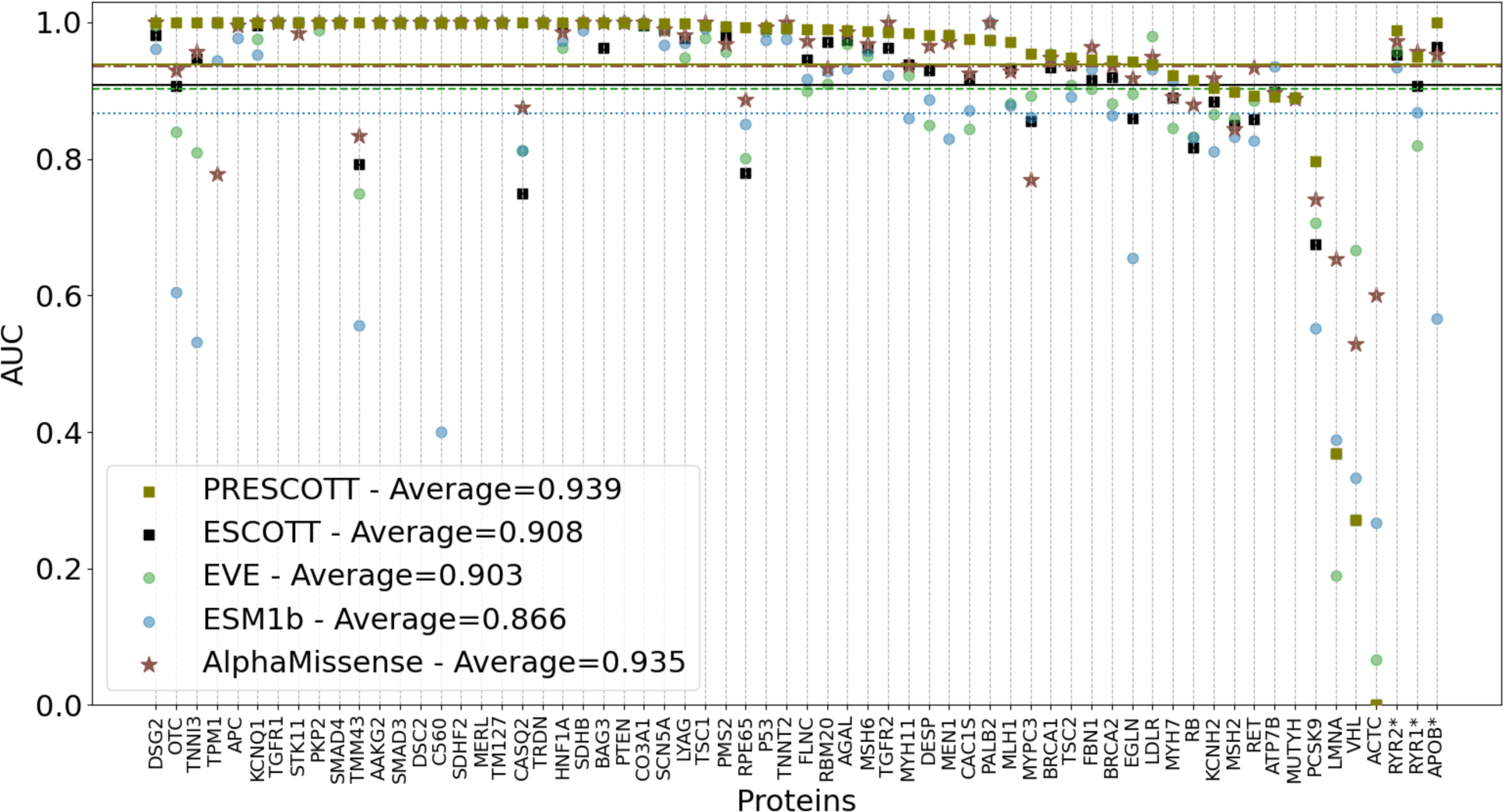
ACMG dataset analysis and comparison of PRESCOTT, ESCOTT, EVE and ESM1b for ACMG human proteins. Global comparative analysis, based on AUC scores, on 64 human proteins in the ACMG dataset of ESCOTT, PRESCOTT, EVE, ESM1b and AlphaMissense. For these proteins, where some are considered in Figure 6A, there is at least 1 benign mutation described in ClinVar.

**Figure S8.**
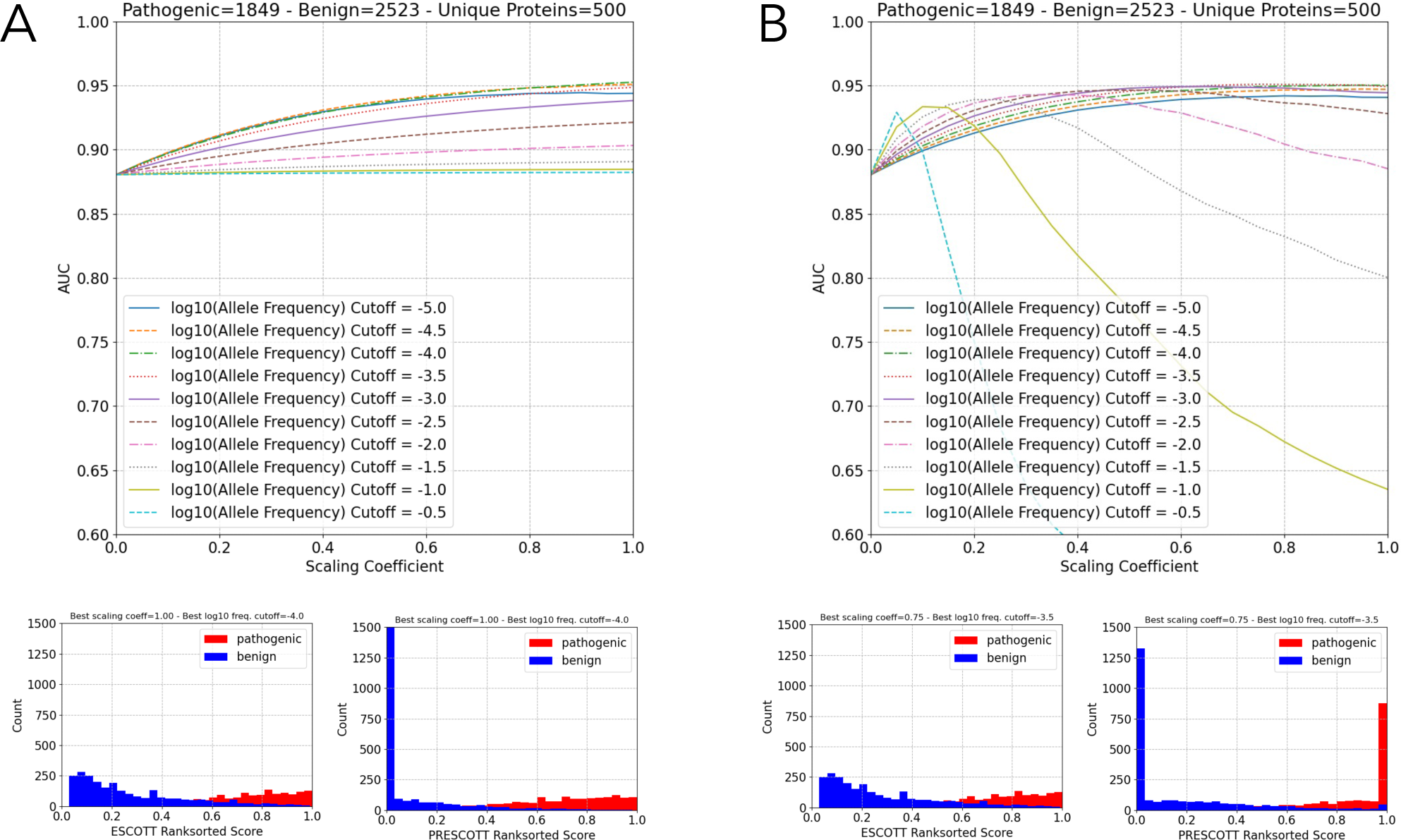
Analysis of the two parameters leading to an optimal PRESCOTT score. The analysis has been realised on 500 proteins containing 1849 pathogenic variants and 2523 benign ones. It is dedicated to fixing the default parameters for allele frequency cut-off and best scaling coefficient. **A.** Algorithm “PRESCOTTscore-low-scores” in Methods. This model improves PRESCOTT scores for highly frequent mutations. Analysis of the allele frequency cut-off parameter (top) leading to a default value of −4 and a best scaling coefficient of 1. The distribution of ESCOTT (bottom left) and PRESCOTT (bottom right) scores is reported for pathogenic and benign variants in the 500 proteins. **B.** Algorithm “PRESCOTTscore-low-and-high-scores” in Methods. This model improves PRESCOTT scores for both highly and lowly frequent mutations. The analysis of the allele frequency cut-off parameter (top) leads to a default value of −3.5 and a best scaling coefficient of 0.75. The distribution of ESCOTT (bottom left; same plot as in A) and PRESCOTT (bottom right) scores is reported for pathogenic and benign variants in the 500 proteins.

**Figure S9.**
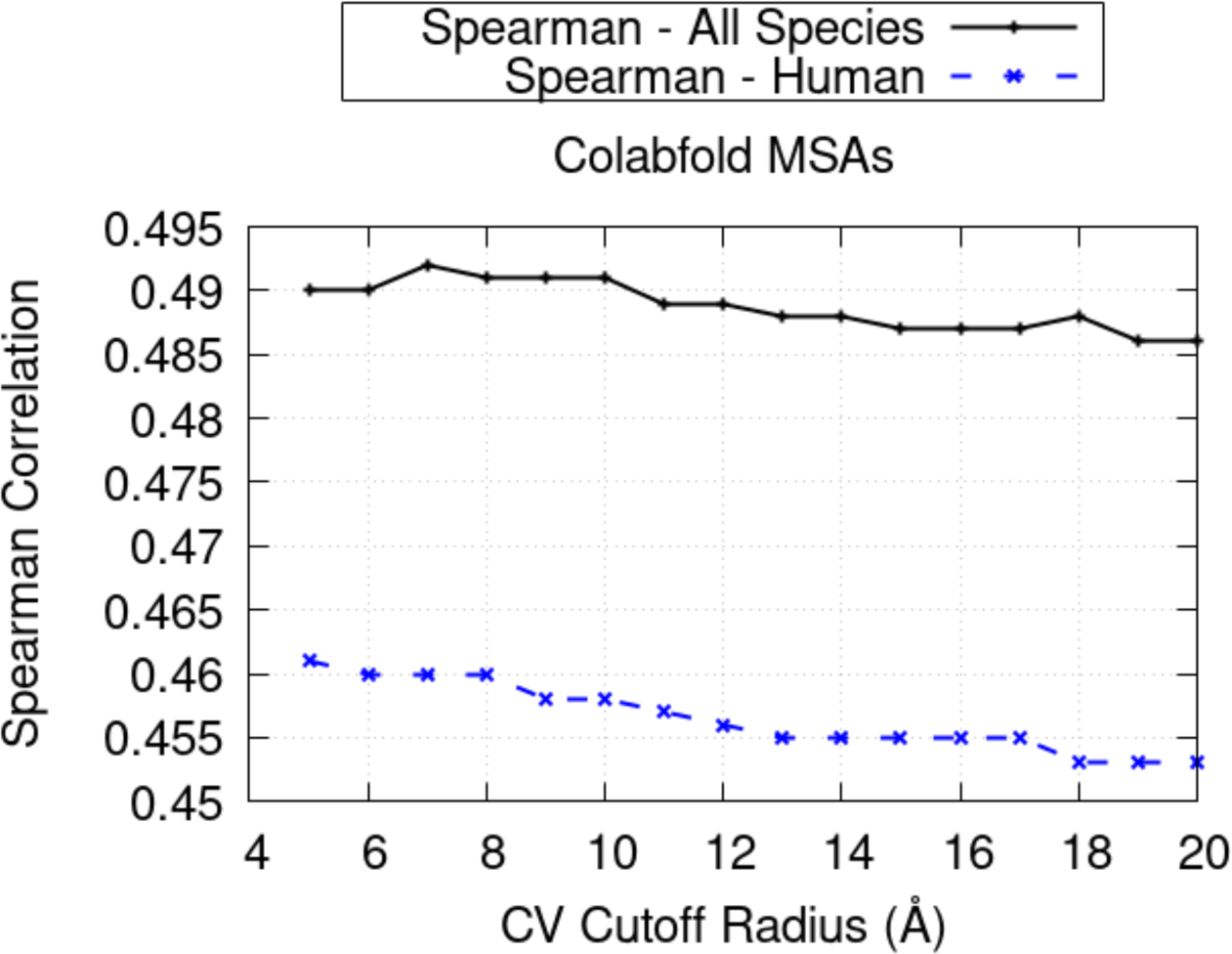
Analysis of ESCOTT performance based on the radius parameter used in the definition of CV. Spearman correlation coefficients computed for the entire set of DMS experiments (solid black line) and the human DMS experiments (dashed blue line) by varying values of the radius that defines the CV formula (x-axis).

**Figure S10.**
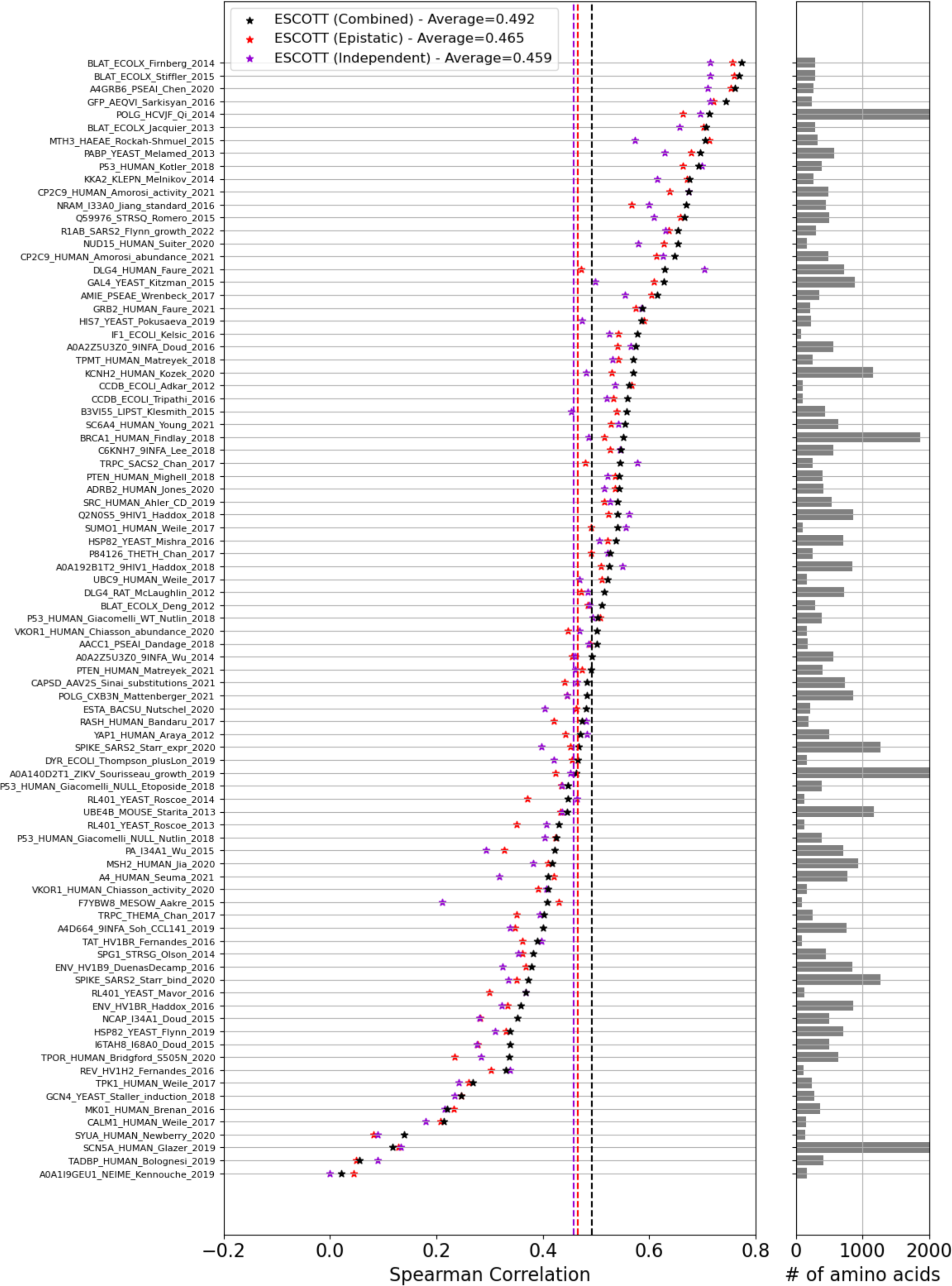
Analysis of the contribution of the independent and epistatic terms in ESCOTT. Stars indicate the average Spearman correlation coefficient between predictions obtained with ESCOTT (black), the ESCOTT independent term only (red) and the ESCOTT epistatic term only (purple) on the 87 experimental data of the ProteinGym dataset. Dashed lines report averages of the three models over the full dataset. Horizontal bars on the right show the number of amino acids in each protein.

**Figure S11.**
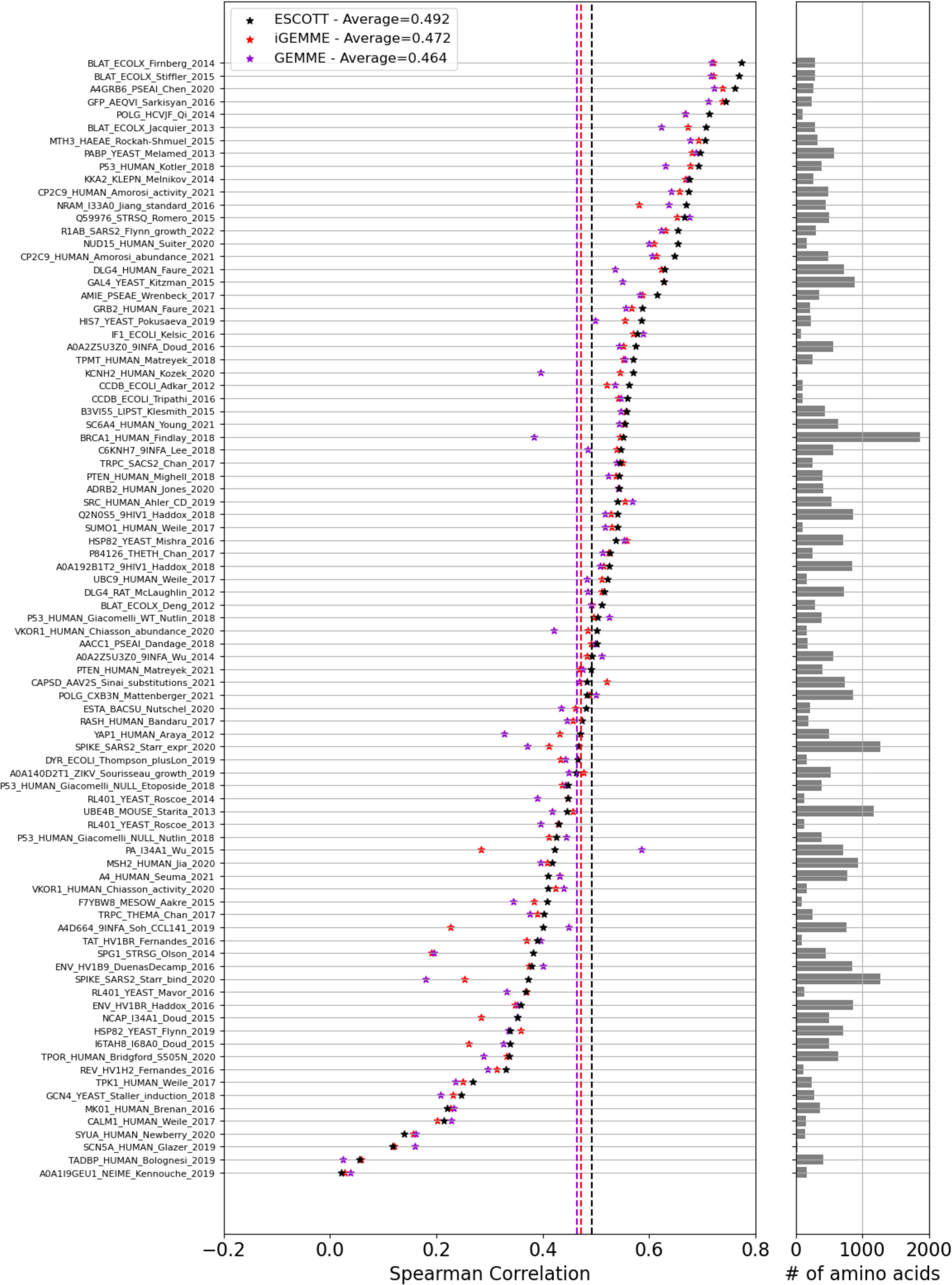
Comparison of ESCOTT, iGEMME and GEMME. Stars indicate the average Spearman correlation coefficient between predictions obtained with ESCOTT (black), iGEMME (red) and GEMME (purple) and the 87 experimental data of the ProteinGym dataset. Dashed lines indicate averages over the full dataset. Horizontal bars on the right show the number of amino acids in each protein.

**Figure S12.**
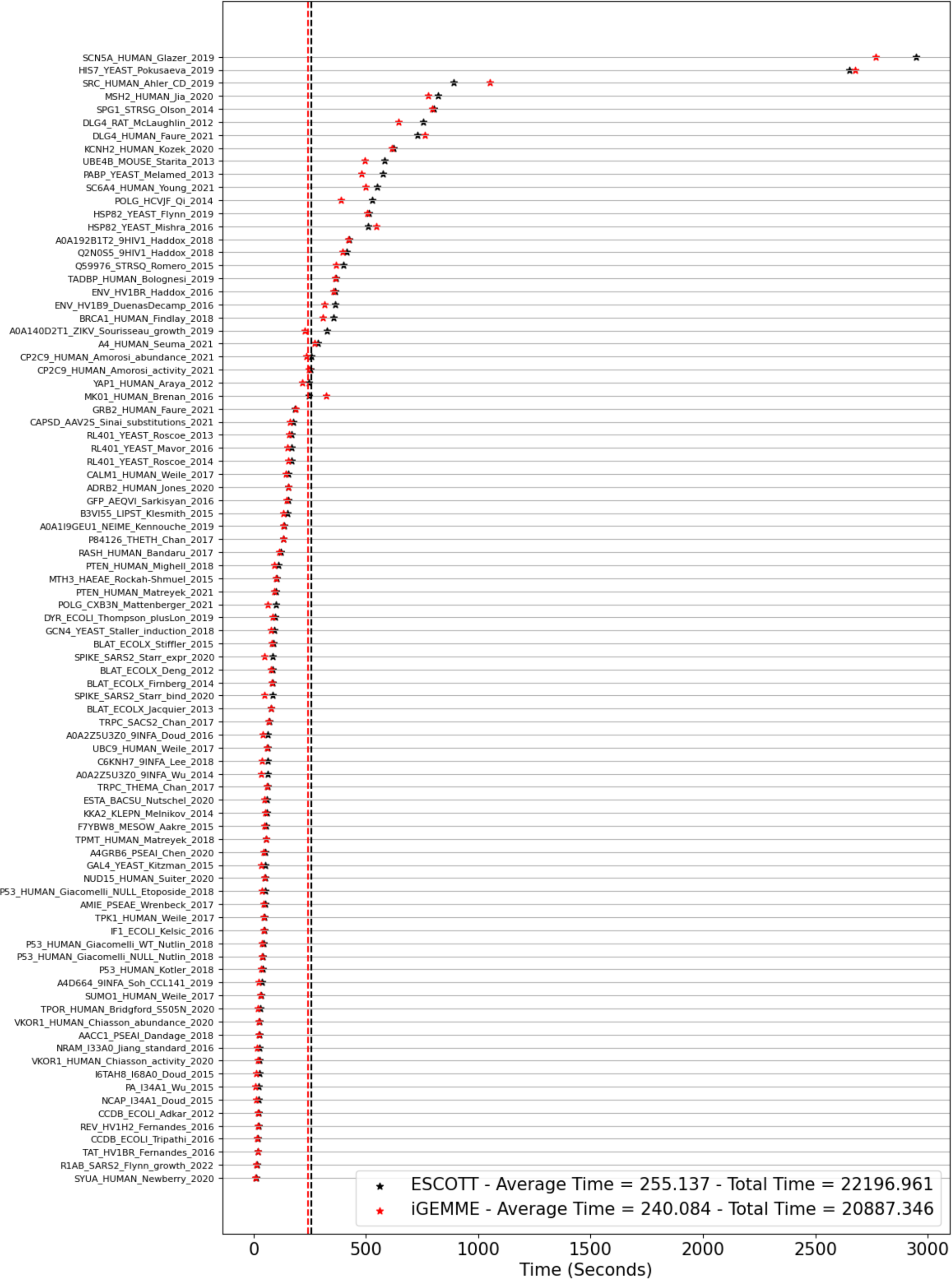
Computation time (in seconds) for all of the experiments in the ProteinGym dataset. The dashed lines indicate the average computing times for ESCOTT=320.531 secs (black) and iGEMME=268.784 secs (red) over 87 experiments.

## REFERENCES

1. UniProt: the Universal Protein knowledgebase in 2023. Nucleic Acids Research. 2023;51(D1):D523–31.

2. Rauer C, Sen N, Waman VP, Abbasian M, Orengo CA. Computational approaches to predict protein functional families and functional sites. Current Opinion in Structural Biology. 2021;70:108–22.

3. Wood V, Lock A, Harris MA, Rutherford K, Bähler J, Oliver SG. Hidden in plain sight: what remains to be discovered in the eukaryotic proteome? Open biology. 2019;9(2):180241.

4. Kustatscher G, Collins T, Gingras AC, Guo T, Hermjakob H, Ideker T, et al. Understudied proteins: opportunities and challenges for functional proteomics. Nature Methods. 2022;19(7):774–9.

5. Vanni C, Schechter MS, Acinas SG, Barberán A, Buttigieg PL, Casamayor EO, et al. Unifying the known and unknown microbial coding sequence space. Elife. 2022;11:e67667.

6. Sinha S, Eisenhaber B, Jensen LJ, Kalbuaji B, Eisenhaber F. Darkness in the human gene and protein function space: widely modest or absent illumination by the life science literature and the trend for fewer protein function discoveries since 2000. Proteomics. 2018;18(21–22):1800093.

7. Stoeger T, Gerlach M, Morimoto RI, Nunes Amaral LA. Large-scale investigation of the reasons why potentially important genes are ignored. PLoS biology. 2018;16(9):e2006643.

8. Oprea TI, Bologa CG, Brunak S, Campbell A, Gan GN, Gaulton A, et al. Unexplored therapeutic opportunities in the human genome. Nature reviews Drug discovery. 2018;17(5):317–32.

9. Wu Y, Liu H, Li R, Sun S, Weile J, Roth FP. Improved pathogenicity prediction for rare human missense variants. The American Journal of Human Genetics. 2021;108(10):1891–906.

10. Hopf TA, Ingraham JB, Poelwijk FJ, Schärfe CP, Springer M, Sander C, et al. Mutation effects predicted from sequence co-variation. Nature biotechnology. 2017;35(2):128–35.

11. Trinquier J, Uguzzoni G, Pagnani A, Zamponi F, Weigt M. Efficient generative modeling of protein sequences using simple autoregressive models. Nature communications. 2021;12(1):5800.

12. Notin P, Dias M, Frazer J, Hurtado JM, Gomez AN, Marks D, et al. Tranception: protein fitness prediction with autoregressive transformers and inference-time retrieval. In: International Conference on Machine Learning. PMLR; 2022. p. 16990–7017.

13. Frazer J, Notin P, Dias M, Gomez A, Min JK, Brock K, et al. Disease variant prediction with deep generative models of evolutionary data. Nature. 2021;599(7883):91–5.

14. Notin P, Van Niekerk L, Kollasch AW, Ritter D, Gal Y, Marks DS. TranceptEVE: Combining family-specific and family-agnostic models of protein sequences for improved fitness prediction. bioRxiv. 2022;2022–12.

15. Marquet C, Heinzinger M, Olenyi T, Dallago C, Erckert K, Bernhofer M, et al. Embeddings from protein language models predict conservation and variant effects. Human genetics. 2022;141(10):1629–47.

16. Riesselman AJ, Ingraham JB, Marks DS. Deep generative models of genetic variation capture the effects of mutations. Nature methods. 2018;15(10):816–22.

17. Meier J, Rao R, Verkuil R, Liu J, Sercu T, Rives A. Language models enable zero-shot prediction of the effects of mutations on protein function. Advances in Neural Information Processing Systems. 2021;34:29287–303.

18. Brandes N, Goldman G, Wang CH, Ye CJ, Ntranos V. Genome-wide prediction of disease variant effects with a deep protein language model. Nature Genetics. 2023;1–11.

19. Cheng J, Novati G, Pan J, Bycroft C, Žemgulytė A, Applebaum T, et al. Accurate proteome-wide missense variant effect prediction with AlphaMissense. Science. 2023 Sep 22;381(6664):eadg7492.

20. Laine E, Karami Y, Carbone A. GEMME: a simple and fast global epistatic model predicting mutational effects. Molecular biology and evolution. 2019;36(11):2604–19.

21. Jumper J, Evans R, Pritzel A, Green T, Figurnov M, Ronneberger O, et al. Highly accurate protein structure prediction with AlphaFold. Nature. 2021;596(7873):583–9.

22. Binder JL, Berendzen J, Stevens AO, He Y, Wang J, Dokholyan NV, et al. AlphaFold illuminates half of the dark human proteins. Current Opinion in Structural Biology. 2022;74:102372.

23. Engelen S, Trojan LA, Sacquin-Mora S, Lavery R, Carbone A. Joint evolutionary trees: a large-scale method to predict protein interfaces based on sequence sampling. PLoS computational biology. 2009;5(1):e1000267.

24. Laine E, Carbone A. Local geometry and evolutionary conservation of protein surfaces reveal the multiple recognition patches in protein-protein interactions. PLoS computational biology. 2015;11(12):e1004580.

25. Wang Z, Moult J. SNPs, protein structure, and disease. Human mutation. 2001;17(4):263–70.

26. Ceres N, Pasi M, Lavery R. A protein solvation model based on residue burial. Journal of Chemical Theory and Computation. 2012;8(6):2141–4.

27. David A, Sternberg MJ. The contribution of missense mutations in core and rim residues of protein– protein interfaces to human disease. Journal of molecular biology. 2015;427(17):2886–98.

28. David A, Razali R, Wass MN, Sternberg MJ. Protein–protein interaction sites are hot spots for disease-associated nonsynonymous SNPs. Human mutation. 2012;33(2):359–63.

29. Gonzalez MW, Kann MG. Chapter 4: Protein interactions and disease. PLoS computational biology. 2012;8(12):e1002819.

30. Negi SS, Braun W. Statistical analysis of physical-chemical properties and prediction of protein-protein interfaces. Journal of molecular modeling. 2007;13:1157–67.

31. Karczewski K, Francioli L. The genome aggregation database (gnomAD). MacArthur Lab. 2017;1–10.

32. Gudmundsson S, Singer-Berk M, Watts NA, Phu W, Goodrich JK, Solomonson M, et al. Variant interpretation using population databases: Lessons from gnomAD. Human mutation. 2022;43(8):1012– 30.

33. Rao RM, Liu J, Verkuil R, Meier J, Canny J, Abbeel P, et al. MSA transformer. In: International Conference on Machine Learning. PMLR; 2021. p. 8844–56.

34. Nijkamp E, Ruffolo J, Weinstein EN, Naik N, Madani A. Progen2: exploring the boundaries of protein language models. arXiv preprint arXiv:220613517. 2022;

35. Hesslow D, Zanichelli N, Notin P, Poli I, Marks D. Rita: a study on scaling up generative protein sequence models. arXiv preprint arXiv:220505789. 2022;

36. Praljak N, Ferguson A. Auto-regressive WaveNet Variational Autoencoders for Alignment-free Generative Protein Design and Fitness Prediction. In: ICLR 2022 Machine Learning for Drug Discovery.

37. Rao R, Meier J, Sercu T, Ovchinnikov S, Rives A. Transformer protein language models are unsupervised structure learners. Biorxiv. 2020;2020–12.

38. Biswas S, Khimulya G, Alley EC, Esvelt KM, Church GM. Low-N protein engineering with data-efficient deep learning. Nature methods. 2021;18(4):389–96.

39. Ferruz N, Schmidt S, Höcker B. ProtGPT2 is a deep unsupervised language model for protein design. Nature communications. 2022;13(1):4348.

40. Alley EC, Khimulya G, Biswas S, AlQuraishi M, Church GM. Unified rational protein engineering with sequence-based deep representation learning. Nature methods. 2019;16(12):1315–22.

41. Karczewski KJ, Francioli LC, Tiao G, Cummings BB, Alföldi J, Wang Q, et al. The mutational constraint spectrum quantified from variation in 141,456 humans. Nature. 2020;581(7809):434–43.

42. Miller DT, Lee K, Abul-Husn NS, Amendola LM, Brothers K, Chung WK, et al. ACMG SF v3. 1 list for reporting of secondary findings in clinical exome and genome sequencing: A policy statement of the American College of Medical Genetics and Genomics (ACMG). Genetics in Medicine. Elsevier; 2022.

43. Mirdita M, von den Driesch L, Galiez C, Martin MJ, S”oding J, Steinegger M. Uniclust databases of clustered and deeply annotated protein sequences and alignments. Nucleic Acids Res. 2017;45(D1):D170–6.

44. Mirdita M, Steinegger M, S”oding J. MMseqs2 desktop and local web server app for fast, interactive sequence searches. Bioinformatics. 2019;35(16):2856–8.

45. Mitchell AL, Almeida A, Beracochea M, Boland M, Burgin J, Cochrane G, et al. MGnify: the microbiome analysis resource in 2020. Nucleic Acids Res. 2019;

46. Mirdita M, Schütze K, Moriwaki Y, Heo L, Ovchinnikov S, Steinegger M. ColabFold: Making Protein folding accessible to all. Nature Methods. 2022;

47. Jumper J, Evans R, Pritzel A, Green T, Figurnov M, Ronneberger O, et al. Highly accurate protein structure prediction with AlphaFold. Nature. 2021;596(7873):583–9.

48. Perica T, Kondo Y, Tiwari SP, McLaughlin SH, Kemplen KR, Zhang X, et al. Evolution of oligomeric state through allosteric pathways that mimic ligand binding. Science. 2014 Dec 19;346(6216):1254346.

49. Marsh JA, Teichmann SA. Predicting pathogenic protein variants. Science. 2023 Sep 22;381(6664):1284–5.

50. Samocha KE, Kosmicki JA, Karczewski KJ, O’Donnell-Luria AH, Pierce-Hoffman E, MacArthur DG, et al. Regional missense constraint improves variant deleteriousness prediction. BioRxiv. 2017;148353.

51. Havrilla JM, Pedersen BS, Layer RM, Quinlan AR. A map of constrained coding regions in the human genome. Nature genetics. 2019;51(1):88–95.

52. Gao H, Hamp T, Ede J, Schraiber JG, McRae J, Singer-Berk M, et al. The landscape of tolerated genetic variation in humans and primates. Science. 2023 Jun 2;380(6648):eabn8153.

53. Fowler DM, Fields S. Deep mutational scanning: a new style of protein science. Nature methods. 2014;11(8):801–7.

54. Hietpas R, Roscoe B, Jiang L, Bolon DN. Fitness analyses of all possible point mutations for regions of genes in yeast. Nature protocols. 2012;7(7):1382–96.

55. Varadi M, Anyango S, Deshpande M, Nair S, Natassia C, Yordanova G, et al. AlphaFold Protein Structure Database: massively expanding the structural coverage of protein-sequence space with high-accuracy models. Nucleic acids research. 2022;50(D1):D439–44.

56. Suzek BE, Wang Y, Huang H, McGarvey PB, Wu CH, Consortium U. UniRef clusters: a comprehensive and scalable alternative for improving sequence similarity searches. Bioinformatics. 2015;31(6):926–32.

57. Steinegger M, Söding J. Clustering huge protein sequence sets in linear time. Nature communications. 2018;9(1):2542.

58. Steinegger M, Mirdita M, Söding J. Protein-level assembly increases protein sequence recovery from metagenomic samples manyfold. Nature methods. 2019;16(7):603–6.

59. Richardson L, Allen B, Baldi G, Beracochea M, Bileschi ML, Burdett T, et al. MGnify: the microbiome sequence data analysis resource in 2023. Nucleic Acids Research. 2023;51(D1):D753–9.

60. Frazer J, Notin P, Dias M, Gomez A, Min JK, Brock K, et al. Disease variant prediction with deep generative models of evolutionary data. Nature. 2021;599(7883):91–5.

61. Mirdita M, Von Den Driesch L, Galiez C, Martin MJ, Söding J, Steinegger M. Uniclust databases of clustered and deeply annotated protein sequences and alignments. Nucleic acids research. 2017;45(D1):D170–6.

62. Mirdita M, Steinegger M, Söding J. MMseqs2 desktop and local web server app for fast, interactive sequence searches. Bioinformatics. 2019;35(16):2856–8.

63. Mitchell AL, Almeida A, Beracochea M, Boland M, Burgin J, Cochrane G, et al. MGnify: the microbiome analysis resource in 2020. Nucleic acids research. 2020;48(D1):D570–8.

64. Mirdita M, Schütze K, Moriwaki Y, Heo L, Ovchinnikov S, Steinegger M. ColabFold: making protein folding accessible to all. Nature methods. 2022;19(6):679–82.

65. Milhavet F, Cuisset L, Hoffman HM, Slim R, El-Shanti H, Aksentijevich I, et al. The infevers autoinflammatory mutation online registry: update with new genes and functions. Hum Mutat. 2008 Jun;29(6):803–8.

66. Richards S, Aziz N, Bale S, Bick D, Das S, Gastier-Foster J, et al. Standards and guidelines for the interpretation of sequence variants: a joint consensus recommendation of the American College of Medical Genetics and Genomics and the Association for Molecular Pathology. Genetics in medicine. 2015;17(5):405–23.

67. Landrum MJ, Lee JM, Benson M, Brown G, Chao C, Chitipiralla S, et al. ClinVar: public archive of interpretations of clinically relevant variants. Nucleic acids research. 2016;44(D1):D862–8.

68. Landrum MJ, Chitipiralla S, Brown GR, Chen C, Gu B, Hart J, et al. ClinVar: improvements to accessing data. Nucleic acids research. 2020;48(D1):D835–44.

69. Kunzmann P, Hamacher K. Biotite: a unifying open source computational biology framework in Python. BMC bioinformatics. 2018;19(1):1–8.

70. Kabsch W, Sander C. Dictionary of protein secondary structure: pattern recognition of hydrogen-bonded and geometrical features. Biopolymers: Original Research on Biomolecules. 1983;22(12):2577–637.

71. Mezei M. A new method for mapping macromolecular topography. Journal of Molecular Graphics and Modelling. 2003;21(5):463–72.

72. Altschul SF, Madden TL, Schäffer AA, Zhang J, Zhang Z, Miller W, et al. Gapped BLAST and PSI-BLAST: a new generation of protein database search programs. Nucleic acids research. 1997;25(17):3389–402.

73. Schäffer AA, Aravind L, Madden TL, Shavirin S, Spouge JL, Wolf YI, et al. Improving the accuracy of PSI-BLAST protein database searches with composition-based statistics and other refinements. Nucleic acids research. 2001;29(14):2994–3005.

74. Pokusaeva VO, Usmanova DR, Putintseva EV, Espinar L, Sarkisyan KS, Mishin AS, et al. An experimental assay of the interactions of amino acids from orthologous sequences shaping a complex fitness landscape. PLoS genetics. 2019;15(4):e1008079.

75. Truong Jr TF, Bepler T. PoET: A generative model of protein families as sequences-of-sequences [Internet]. arXiv; 2023 [cited 2023 Sep 28]. Available from: http://arxiv.org/abs/2306.06156

76. Schrodinger LLC. The PyMOL molecular graphics system. Version. 2015;1:8.

